# Human immunodeficiency virus and tuberculosis coinfection prevalence and treatment outcomes in Cameroon: a systematic review and meta-analysis

**DOI:** 10.64898/2026.11.07.26367493

**Authors:** Fabrice Zobel Lekeumo Cheuyem, Rick Tchamani, Arole Darwin Touko, Chabeja Achangwa, Clovis Josué Kachiwouo Mapouo, Germerie Campbell Manfouo Fouodjeu, Ariane Nouko, Saidou Modibo

**Author notes:** Corresponding author’s address: Fabrice Zobel Lekeumo Cheuyem.

## Abstract

**Background:** Human immunodeficiency virus (HIV) and tuberculosis (TB) form one of the deadliest syndemics, and sub-Saharan Africa bears over 80% of HIV-associated TB deaths. In Cameroon, evidence on HIV/TB coinfection remains unsynthesized across regions and populations, obscuring the national burden. Hence, we estimated the prevalence of HIV/TB coinfection and treatment outcomes in Cameroon.

**Methods:** Following PRISMA 2020 (PROSPERO: CRD420261399169), we searched PubMed, Scopus, Embase, Web of Science, Cochrane Library, African Journals Online, and grey literature for English- and French-language studies conducted in Cameroon. Reports were independently screened, their data extracted, and their methodology appraised using Joanna Briggs Institute tools. Random-effects models pooled prevalence, treatment outcomes, and adjusted odds ratios. Heterogeneity was assessed with *I*².

**Results:** Seventy studies (1995–2023) were included, 87.1% at low risk of bias. Pooled HIV prevalence among TB patients was 29.2% (95% CI: 26.3–32.3; 67 studies; n=57,169; *I*² = 97.8%), with no significant evidence of publication bias. Prevalence was higher from 2007 onwards (29.9%; 62 studies; n=55,358) than before (20.5%; 5 studies; n=1,811). It was highest in the North-West (41.9%; 7 studies; n=6,324) versus Littoral (25.0%; 15 studies; n=10,419) and West (23.8%; 6 studies; n=4,206); and in extrapulmonary TB (42.3%; 3 studies; n=371) versus mixed forms (37.7%; 21 studies; n=37,239) and pulmonary TB (23.9%; 37 studies; n=16,096) (all *p*<0.05). Among coinfected patients, pooled treatment success was 75.9% (95% CI: 70.4–80.8; 8 studies; n=4,230), mortality 21.1% (95% CI: 11.9–34.7; 10 studies; n=4,403), loss to follow-up 7.7% (95% CI: 4.3–13.3; 5 studies; n=3,103), and treatment failure 0.3% (95% CI: 0.1–1.5; 6 studies; n=3,520). Lack of antiretroviral therapy (ART) increased the odds of unfavorable outcomes (AOR = 1.90; 95% CI: 1.15–3.12; 2 studies).

**Conclusions:** Approximately three in ten TB patients in Cameroon are coinfected with HIV, with marked regional and clinical variation. Treatment success remains below national targets. Strengthening TB-HIV service integration, routine HIV testing, and timely ART initiation and continuation is essential to reduce the HIV-associated TB morbidity and mortality in the country.

## 1. Background

Human immunodeficiency virus (HIV) and *Mycobacterium tuberculosis* (TB) constitute one of the most lethal syndemic associations in infectious diseases [1]. HIV/TB coinfection represents a major syndemic interaction in which HIV impairs cell-mediated immunity, promoting reactivation and rapid progression of tuberculosis, while TB infection induces chronic immune activation that may accelerate viral replication and HIV progression due to profound impairment of cell-mediated immunity [2, 3]. Co-infected patients generally exhibit higher HIV viral loads, larger viral reservoirs, more pronounced immune activation, as well as a higher frequency of disseminated tuberculosis [4]. A recent global systematic review and meta-analysis reported substantial geographical variation in the prevalence of HIV/TB coinfection, with the highest pooled prevalence observed in Africa (22%), followed by North America (21%), South America (16%), Europe (10%), and Asia (7%). Meta-regression analyses further identified male sex, lower educational attainment, and drug-resistant tuberculosis as factors significantly associated with higher HIV–TB coinfection prevalence [5].

Recent reports revealed that HIV-associated TB cases accounted for 6.3% of all new TB cases in 2022, with approximately 167,000 deaths attributed to TB among people living with HIV worldwide. In the same year, an estimated 671,000 people living with HIV developed TB, with the highest burden occurring in the WHO African Region [6]. The intersection of the two epidemics remains a major public health concern in sub-Saharan Africa, which bears a disproportionate share of the global burden, accounting for more than 80% of HIV-associated TB deaths [7]. HIV-TB prevalence also varies considerably across sub-Saharan Africa, with prevalence estimates of 43.7%, 41.3%, 31.1%, and 25.5% in Southern, Central, Eastern, and Western sub-Saharan Africa, respectively [8]. Moreover, HIV/TB coinfection is associated with a substantial mortality burden among children in sub- Saharan African countries, with a pooled mortality estimate of 12.96%. Several clinical and treatment-related factors have been identified as predictors of premature mortality during HIV/TB coinfection, including WHO clinical stages III and IV, poor or fair adherence to antiretroviral therapy (ART), missed isoniazid preventive therapy, hemoglobin levels ≤10 g/dL, CD4 counts below the recommended threshold, and absence of cotrimoxazole preventive therapy (CPT) [9].

In Cameroon, HIV/TB coinfection is a major public health problem. Studies published in 2024 have further highlighted the substantial burden of HIV/TB coinfection in different regions of Cameroon, with reported coinfection rates of 47.7% in the North-West Region, 24.1% in the Littoral Region, and 40.2% in the South-West Region [1–3]. These findings suggest considerable regional variation in the magnitude of HIV–TB coinfection and underscore the persistent overlap between the two epidemics in Cameroon. Studies conducted in Cameroon have also identified several factors associated with the occurrence of TB among people living with HIV, including anemia, tobacco smoking, alcohol consumption, detectable HIV viral load, CD4 count ≤200 cells/µL, and sex, which were significantly associated with the development of TB among people living with HIV [10].

However, the available evidence on HIV–TB coinfection in Cameroon remains unsynthesized, with studies conducted in specific regions, health facilities, and populations and substantial variation in study designs and reported estimates. Consequently, the overall national burden of HIV/TB coinfection remains difficult to ascertain, and the extent to which regional and population-level differences contribute to the observed variability has not been comprehensively quantified. Moreover, despite the availability of individual studies reporting the burden and associated factors of HIV/TB coinfection, a comprehensive systematic review and meta-analysis synthesizing the available evidence from Cameroon is lacking. Therefore, this systematic review and meta-analysis aims to estimate the prevalence of HIV/TB coinfection and its treatment outcomes in Cameroon.

## **2.** Methods

### 2.1. Study design

This review was conducted in accordance with the Preferred Reporting Items for Systematic Reviews and Meta-Analyses (PRISMA) 2020 and was registered in the International Prospective Register of Systematic Reviews under reference CRD420261399169 [11].

### 2.2. Eligibility criteria

#### Inclusion criteria

This systematic review included observational (cross-sectional, cohort, and case–control) and interventional studies with quantitative outcomes conducted in Cameroon. Studies were eligible if they reported data sufficient to estimate HIV/TB co- infection prevalence, with no restriction on study timeframe. Eligible studies included subjects of any age with bacteriologically confirmed or clinically diagnosed tuberculosis. Studies published in either English or French were considered for inclusion.

#### Exclusion criteria

Studies were excluded if they were reviews (narrative, systematic, or meta-analyses), editorials, commentaries, opinion pieces, case reports, conference abstracts, or letters without primary data. Also excluded were studies conducted outside Cameroon; those that did not report relevant outcomes related to HIV/TB co-infection; those with insufficient methodological information, incomplete data, or corrupted or unextractable outcome data; animal studies and in vitro or laboratory-only investigations. Finally, duplicate publications of the same dataset were excluded, with the most comprehensive and recent version retained for analysis.

### 2.3. Search strategy

#### Databases

To identify all relevant literature for this review, we conducted a comprehensive search across several of databases and other sources. Our search strategy included major scientific databases such as PubMed, Scopus, Embase, Web of Science, and Cochrane Library. To ensure we captured research from Cameroonian scholars and journals, we also conducted searches on African Journals Online (AJOL). In addition to these databases, we searched Google Scholar for grey literature, including unpublished research and preprints. Moreover, we conducted a manual search on the reference lists of included reports. Data searching was concluded on 2026 August 01.

#### Search terms

To effectively capture the relevant literature, we used a comprehensive search strategy that combines Medical Subject Headings (MeSH) terms and Boolean connectors. The search used the following terms and logic: (“HIV” OR “*Human immunodeficiency virus*” OR “AIDS”) AND (“Tuberculosis” OR “TB” OR “*Mycobacterium tuberculosis*”) AND (“Cameroon” OR “Cameroun” OR Cameroonian). The literature search strategy was adapted to suit each database. (**Table S1**). All retrieved articles were initially screened and deduplicated; the remaining records were then assessed against the inclusion criteria. Full texts were then independently screened by two reviewers (FZLC and RT), and any disagreements were resolved through discussion.

### 2.4. Data extraction

Key data points were extracted from each study using a Microsoft Excel spreadsheet to ensure a thorough and consistent analysis. For each study, the following information was recorded: author, year of study completion, study period, study design, region and study setting, study population, sample size (number of TB cases screened for HIV), localization of TB disease, and prevalence of HIV/TB co-infection. Data extraction was performed independently by two study investigators (FZLC and ADT), with discrepancies resolved through discussion to reach consensus or consulting a third reviewer (CA).

### 2.5. Outcomes measurement and operational definition

A patient was considered to have laboratory-confirmed TB if a specimen tested positive bacteriologically or TB-specific real-time PCR. Similarly, a case was defined as laboratory- confirmed HIV if a specimen tested positive on an HIV antigen-specific assay or an HIV- specific real-time PCR. HIV/TB coinfection was defined as the concurrent diagnosis of HIV infection and active tuberculosis [12].

The primary outcome of this review was the prevalence of HIV/TB coinfection, which was calculated by dividing the number of participants with HIV infection by the total number of TB cases screened for HIV. The secondary outcome included the pooled rates of treatment success (cured and treatment completion), loss to follow-up, treatment failure, and mortality among active TB patients coinfected with HIV. Treatment outcomes were defined according to 2020 WHO guidelines [13]. Treatment failure as defined as a patient whose treatment regimen required termination or permanent substitution with a new regimen or treatment strategy. Cure was referred to a patient with pulmonary TB and bacteriologically confirmed TB at treatment initiation who completed treatment as recommended by national policy, with evidence of bacteriological response and no evidence of failure. Treatment completion was defined as a patient who completed treatment as recommended by national policy but whose outcome did not meet the criteria for cure or treatment failure. Mortality referred to a patient who died before starting treatment or during the course of treatment. Lost to follow-up (LTFU) referred to a patient who did not start treatment or whose treatment was interrupted for two consecutive months or more. Treatment success was defined as the sum of all patients cured and those who completed treatment. The prevalence of each outcome was computed by dividing the number of patients experiencing the specific outcome by the total number of treated patients and was expressed as a percentage. Secondary outcomes included factors associated with unfavorable outcomes. Unfavorable outcome included patients declared dead, lost to follow-up, or experiencing treatment failure [12, 14].

### 2.6. Risk of bias assessment

The methodological quality of the included studies was assessed using the Joanna Briggs Institute (JBI) critical appraisal tools, selected according to each study’s design [15]. For cohort studies, the checklist contains eleven items assessing: if the two groups were similar and recruited from the same population; exposures were measured similarly to assign people to exposed and unexposed groups; the exposure was measured in a valid and reliable way; confounding factors were identified and strategies to deal with them stated; whether the groups/participants were free of the outcome at the start of the study (or at the moment of exposure); outcomes were measured in a valid and reliable way; whether the follow-up time was sufficient for outcomes to occur; follow-up was complete, with reasons for loss to follow-up described and explored; strategies to address incomplete follow-up were utilized; and appropriate statistical analysis was used. Each criterion was scored as 1 (yes) or 0 (no or unclear). In this study, we defined and applied the following categorization of the overall risk of bias: low = ≥70%, moderate = 50%-69%, and high risk of bias = <50% of "Yes" scores. The quality assessment was performed independently by two reviewers (FZLC and RT) and any disagreements were resolved by discussion or, if necessary, by involving a third reviewer (CA).

### 2.7. Statistical analysis

The random-effects model was used to pool the prevalence estimates and associated factors. Heterogeneity was assessed using the Higgins’ *I*², with values categorized as low (<25%), moderate (25–75%), or high (>75%). Generalized linear mixed models (GLMM) with probit-logit transformation (PLOGIT) were employed to effectively handle meta-analyses of binary data and accommodate studies with 0% or 100% event rates without requiring continuity corrections [16]. Subgroup analyses were performed for each outcome where sufficient data were available. Statistical significance was set at *p* < 0.05. All analyses were performed using the ’meta’ package in R Statistics version 4.6.1 [17]. Adjusted odds ratios were pooled to identify factors significantly associated with unfavorable outcomes.

### 2.8. Publication bias and sensitivity analysis

Publication bias was assessed graphically through funnel plots and statistically using Egger’s linear regression test and Begg’s rank correlation test, with a two-tailed *p* < 0.05 considered indicative of significant publication bias [18, 19]. In addition, a leave-one-out sensitivity analysis was conducted to evaluate the influence of individual studies on the overall findings.

## **3.** Results

### 3.1. Study selection and inclusion

A systematic search of six electronic databases identified 1,253 records: PubMed (n = 182), Scopus (n = 239), Web of Science (n = 276), Embase (n = 521), the Cochrane Library (n = 17), and African Journals Online (n = 18). After removing 197 duplicates, 1,056 records remained for title and abstract screening. Of these, 984 were excluded, and the full texts of the remaining 72 reports were retrieved and assessed for eligibility. Twenty-six reports were excluded: 10 did not report the outcome of interest, two presented data aggregated across several countries, nine used an incorrect denominator, four contained duplicate findings, and one was a review article. This resulted in 46 studies from the database searches. Searches of other sources identified 3,203 records. Following screening, 31 reports were retrieved and assessed for eligibility, of which seven were excluded because they did not report the outcome of interest. The remaining 24 studies met the inclusion criteria. Overall, 70 studies were included in the systematic review and meta-analysis (**Fig. 1**).

**Fig. 1.**
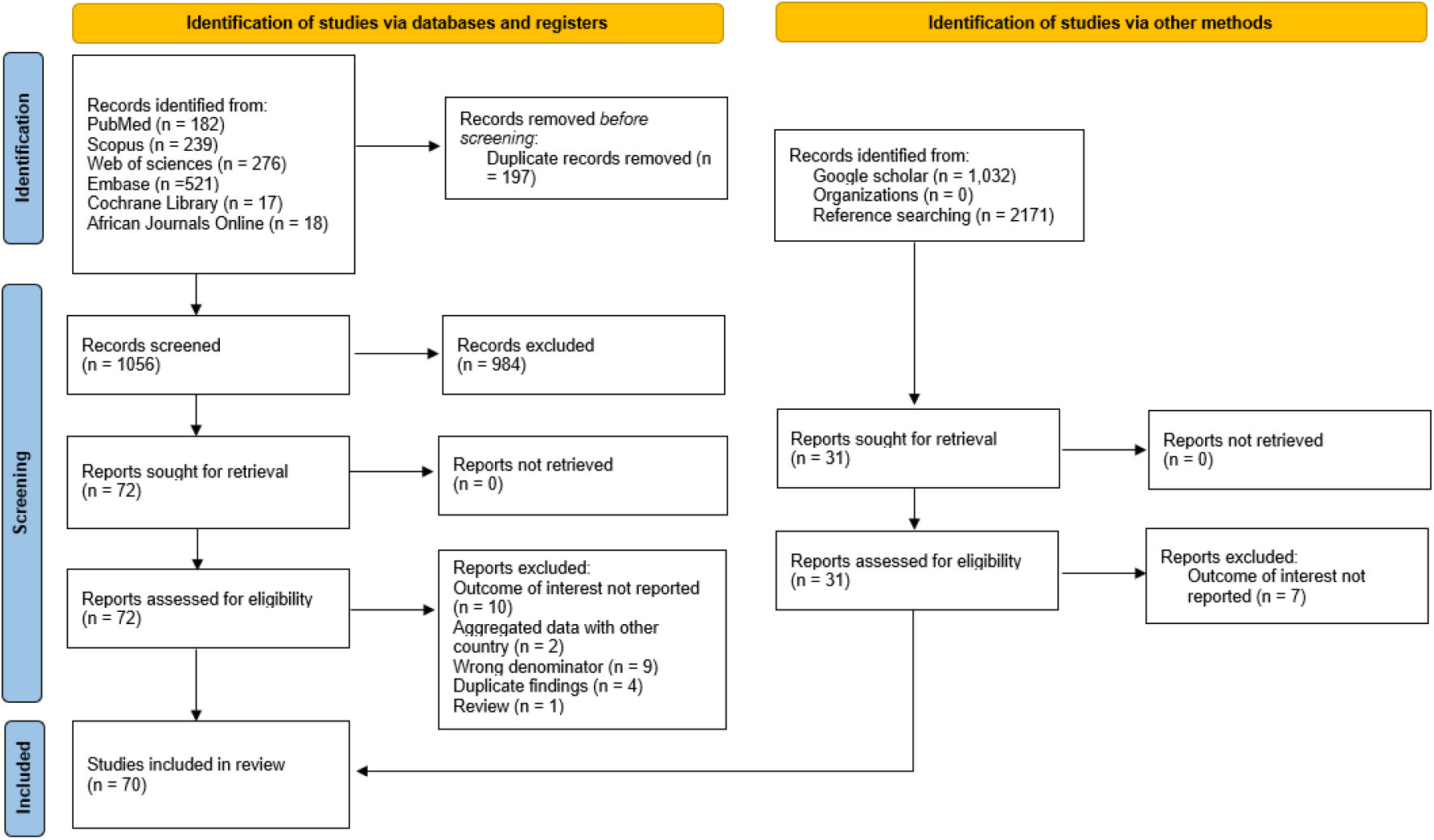
PRISMA diagram flow from study identification to inclusion in the systematic review and meta-analysis [11]

### 3.2. Characteristics of included studies

A comprehensive synthesis of 70 studies conducted in Cameroon between 1995 and 2023, examining HIV/TB coinfection and associated treatment outcomes. The studies were predominantly cross-sectional (n = 45; 64.3%) and cohort designs (n = 22; 31.4%), with two case-control studies (2.9%) and one randomized controlled trial (1.4%), and were distributed across most regions, though heavily concentrated in the Centre (n = 19; 27.1%), Littoral (n = 15; 21.4%), and with 10 studies (14.3%) conducted across multiple regions. The majority focused on adult TB patients (n = 38; 54.3%), with fewer studies targeting retreated patients (n = 3; 4.3%), rifampicin-resistant TB (n = 3; 4.3%), multidrug-resistant TB (n = 3; 4.3%), or pediatric populations (n = 3; 4.3%). Pulmonary TB (PTB) was the most studied clinical form (n = 37; 52.9%), followed by mixed cases (n = 22; 31.4%), while extrapulmonary TB (EPTB) specific studies were limited (n = 4; 5.7%). HIV coinfection prevalence varied widely across studies, ranging from 9.8% to 68.5%. Among the 11 studies (15.7%) reporting treatment outcomes among HIV-positive patients, treatment success ranged from 60.8% to 84.0%, mortality ranged from 6.2% to 29.4%, and loss to follow-up varied from 3.8% to 19.3%. The risk of bias assessment rated the vast majority of studies (n = 61; 87.1%) as low risk, indicating generally sound methodological quality (**Table 1**).

**Table 1.**
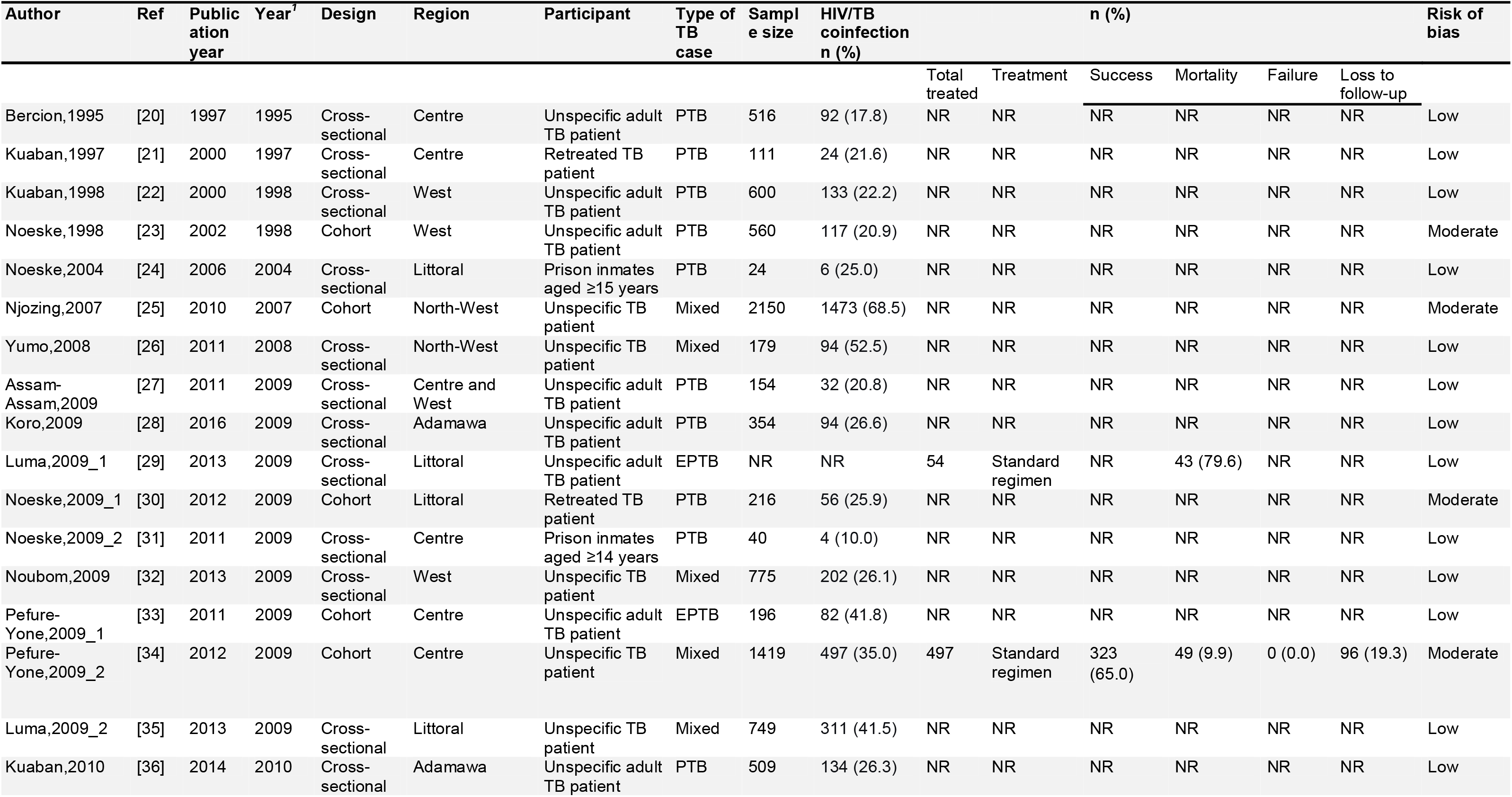
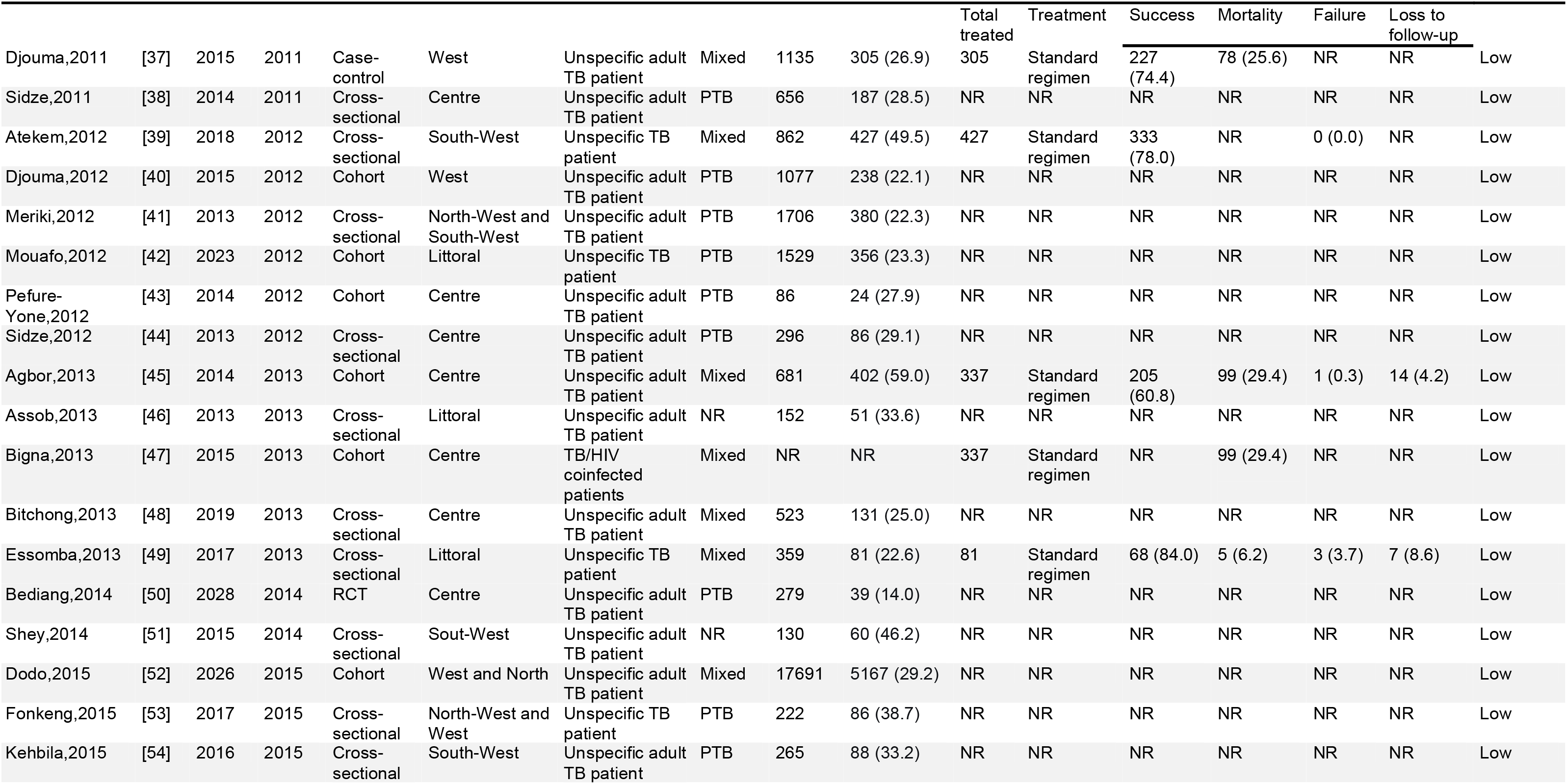
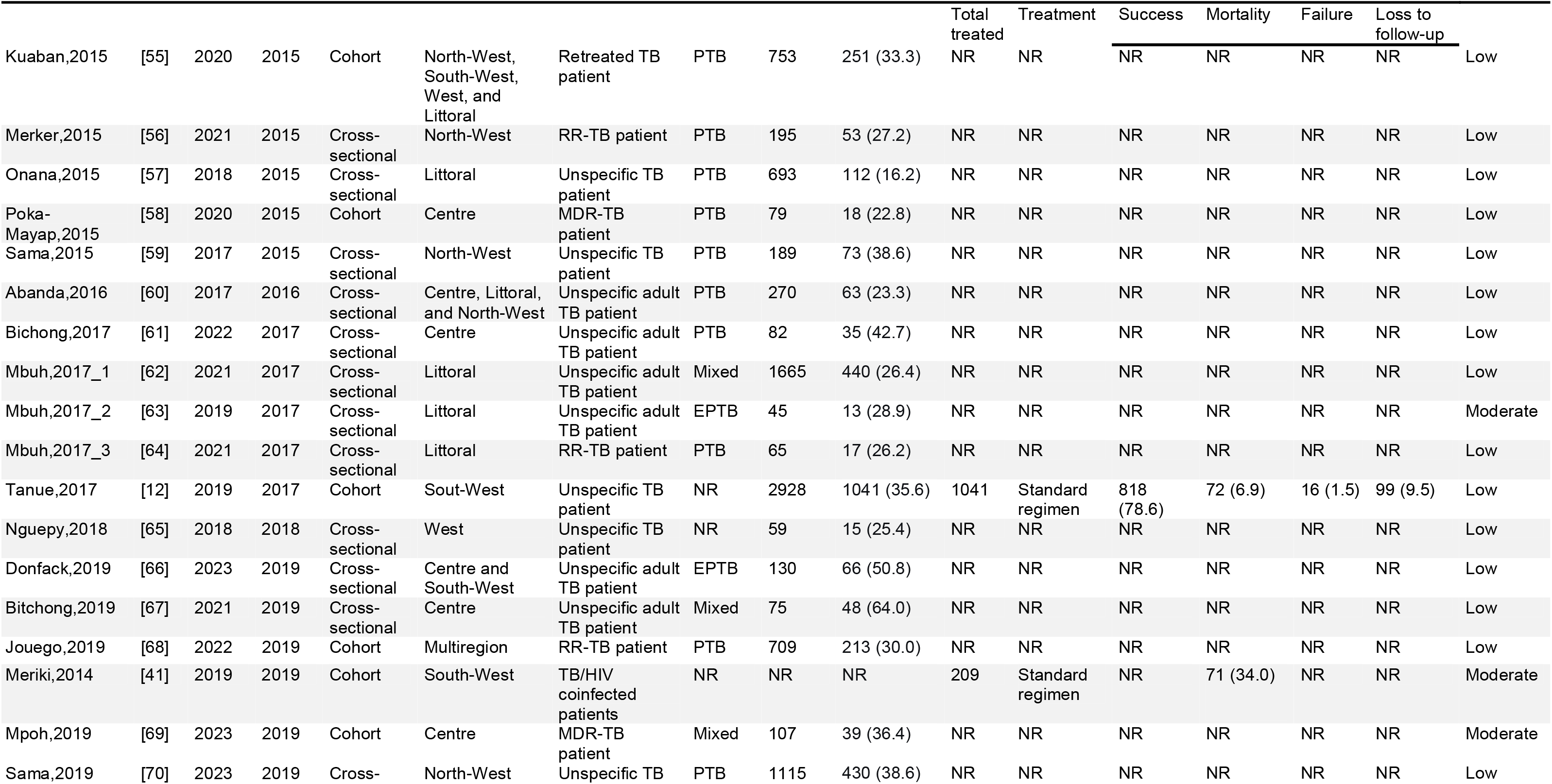
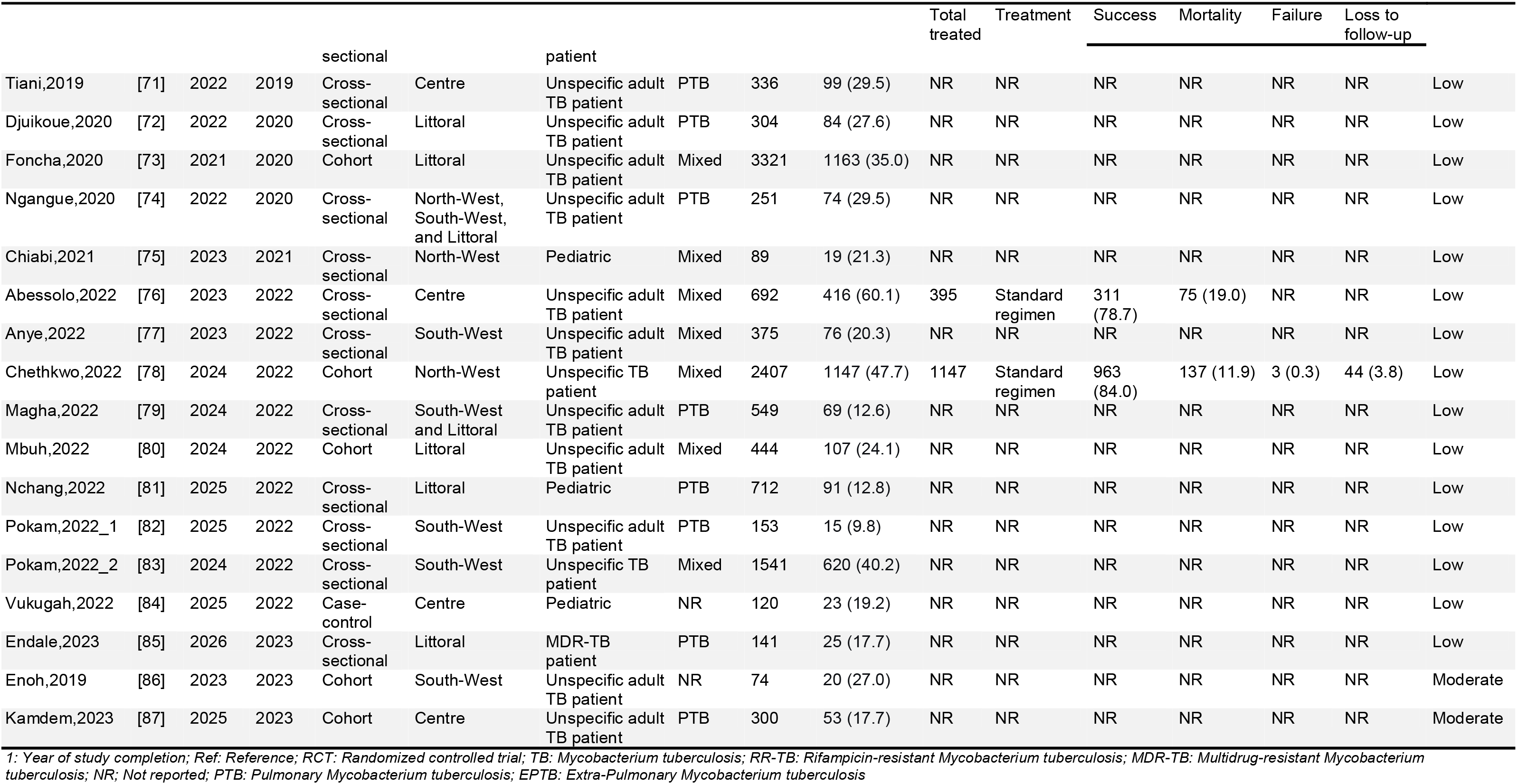
Synthesis of study characteristics

| **Author** | **Ref** | **Publication year** | **Year<sup>1</sup>** | **Design** | **Region** | **Participant** | **Type of TB case** | **Sample size** | **HIV/TB coinfection n (%)** |  |  | **n (%)** |  |  |  | **Risk of bias** |
|  |  |  |  |  |  |  |  |  |  | Total treated | Treatment | Success | Mortality | Failure | Loss to follow-up |  |
|---|---|---|---|---|---|---|---|---|---|---|---|---|---|---|---|---|
| Bercion,1995 | [20] | 1997 | 1995 | Cross-sectional | Centre | Unspecific adult TB patient | PTB | 516 | 92 (17.8) | NR | NR | NR | NR | NR | NR | Low |
| Kuaban,1997 | [21] | 2000 | 1997 | Cross-sectional | Centre | Retreated TB patient | PTB | 111 | 24 (21.6) | NR | NR | NR | NR | NR | NR | Low |
| Kuaban,1998 | [22] | 2000 | 1998 | Cross-sectional | West | Unspecific adult TB patient | PTB | 600 | 133 (22.2) | NR | NR | NR | NR | NR | NR | Low |
| Noeske,1998 | [23] | 2002 | 1998 | Cohort | West | Unspecific adult TB patient | PTB | 560 | 117 (20.9) | NR | NR | NR | NR | NR | NR | Moderate |
| Noeske,2004 | [24] | 2006 | 2004 | Cross-sectional | Littoral | Prison inmates aged ≥15 years | PTB | 24 | 6 (25.0) | NR | NR | NR | NR | NR | NR | Low |
| Njozing,2007 | [25] | 2010 | 2007 | Cohort | North-West | Unspecific TB patient | Mixed | 2150 | 1473 (68.5) | NR | NR | NR | NR | NR | NR | Moderate |
| Yumo,2008 | [26] | 2011 | 2008 | Cross-sectional | North-West | Unspecific TB patient | Mixed | 179 | 94 (52.5) | NR | NR | NR | NR | NR | NR | Low |
| Assam-Assam,2009 | [27] | 2011 | 2009 | Cross-sectional | Centre and West | Unspecific adult TB patient | PTB | 154 | 32 (20.8) | NR | NR | NR | NR | NR | NR | Low |
| Koro,2009 | [28] | 2016 | 2009 | Cross-sectional | Adamawa | Unspecific adult TB patient | PTB | 354 | 94 (26.6) | NR | NR | NR | NR | NR | NR | Low |
| Luma,2009_1 | [29] | 2013 | 2009 | Cross-sectional | Littoral | Unspecific adult TB patient | EPTB | NR | NR | 54 | Standard regimen | NR | 43 (79.6) | NR | NR | Low |
| Noeske,2009_1 | [30] | 2012 | 2009 | Cohort | Littoral | Retreated TB patient | PTB | 216 | 56 (25.9) | NR | NR | NR | NR | NR | NR | Moderate |
| Noeske,2009_2 | [31] | 2011 | 2009 | Cross-sectional | Centre | Prison inmates aged ≥14 years | PTB | 40 | 4 (10.0) | NR | NR | NR | NR | NR | NR | Low |
| Noubom,2009 | [32] | 2013 | 2009 | Cross-sectional | West | Unspecific TB patient | Mixed | 775 | 202 (26.1) | NR | NR | NR | NR | NR | NR | Low |
| Pefure-Yone,2009_1 | [33] | 2011 | 2009 | Cohort | Centre | Unspecific adult TB patient | EPTB | 196 | 82 (41.8) | NR | NR | NR | NR | NR | NR | Low |
| Pefure-Yone,2009_2 | [34] | 2012 | 2009 | Cohort | Centre | Unspecific TB patient | Mixed | 1419 | 497 (35.0) | 497 | Standard regimen | 323 (65.0) | 49 (9.9) | 0 (0.0) | 96 (19.3) | Moderate |
| Luma,2009_2 | [35] | 2013 | 2009 | Cross-sectional | Littoral | Unspecific TB patient | Mixed | 749 | 311 (41.5) | NR | NR | NR | NR | NR | NR | Low |
| Kuaban,2010 | [36] | 2014 | 2010 | Cross-sectional | Adamawa | Unspecific adult TB patient | PTB | 509 | 134 (26.3) | NR | NR | NR | NR | NR | NR | Low |
|  |  |  |  |  |  |  |  |  |  | Total treated | Treatment | Success | Mortality | Failure | Loss to follow-up |  |
| Djouma,2011 | [37] | 2015 | 2011 | Case-control | West | Unspecific adult TB patient | Mixed | 1135 | 305 (26.9) | 305 | Standard regimen | 227 (74.4) | 78 (25.6) | NR | NR | Low |
| Sidze,2011 | [38] | 2014 | 2011 | Cross-sectional | Centre | Unspecific adult TB patient | PTB | 656 | 187 (28.5) | NR | NR | NR | NR | NR | NR | Low |
| Atekem,2012 | [39] | 2018 | 2012 | Cross-sectional | South-West | Unspecific TB patient | Mixed | 862 | 427 (49.5) | 427 | Standard regimen | 333 (78.0) | NR | 0 (0.0) | NR | Low |
| Djouma,2012 | [40] | 2015 | 2012 | Cohort | West | Unspecific adult TB patient | PTB | 1077 | 238 (22.1) | NR | NR | NR | NR | NR | NR | Low |
| Meriki,2012 | [41] | 2013 | 2012 | Cross-sectional | North-West and South-West | Unspecific adult TB patient | PTB | 1706 | 380 (22.3) | NR | NR | NR | NR | NR | NR | Low |
| Mouafo,2012 | [42] | 2023 | 2012 | Cohort | Littoral | Unspecific TB patient | PTB | 1529 | 356 (23.3) | NR | NR | NR | NR | NR | NR | Low |
| Pefure-Yone,2012 | [43] | 2014 | 2012 | Cohort | Centre | Unspecific adult TB patient | PTB | 86 | 24 (27.9) | NR | NR | NR | NR | NR | NR | Low |
| Sidze,2012 | [44] | 2013 | 2012 | Cross-sectional | Centre | Unspecific adult TB patient | PTB | 296 | 86 (29.1) | NR | NR | NR | NR | NR | NR | Low |
| Agbor,2013 | [45] | 2014 | 2013 | Cohort | Centre | Unspecific adult TB patient | Mixed | 681 | 402 (59.0) | 337 | Standard regimen | 205 (60.8) | 99 (29.4) | 1 (0.3) | 14 (4.2) | Low |
| Assob,2013 | [46] | 2013 | 2013 | Cross-sectional | Littoral | Unspecific adult TB patient | NR | 152 | 51 (33.6) | NR | NR | NR | NR | NR | NR | Low |
| Bigna,2013 | [47] | 2015 | 2013 | Cohort | Centre | TB/HIV coinfected patients | Mixed | NR | NR | 337 | Standard regimen | NR | 99 (29.4) | NR | NR | Low |
| Bitchong,2013 | [48] | 2019 | 2013 | Cross-sectional | Centre | Unspecific adult TB patient | Mixed | 523 | 131 (25.0) | NR | NR | NR | NR | NR | NR | Low |
| Essomba,2013 | [49] | 2017 | 2013 | Cross-sectional | Littoral | Unspecific TB patient | Mixed | 359 | 81 (22.6) | 81 | Standard regimen | 68 (84.0) | 5 (6.2) | 3 (3.7) | 7 (8.6) | Low |
| Bediang,2014 | [50] | 2028 | 2014 | RCT | Centre | Unspecific adult TB patient | PTB | 279 | 39 (14.0) | NR | NR | NR | NR | NR | NR | Low |
| Shey,2014 | [51] | 2015 | 2014 | Cross-sectional | Sout-West | Unspecific adult TB patient | NR | 130 | 60 (46.2) | NR | NR | NR | NR | NR | NR | Low |
| Dodo,2015 | [52] | 2026 | 2015 | Cohort | West and North | Unspecific adult TB patient | Mixed | 17691 | 5167 (29.2) | NR | NR | NR | NR | NR | NR | Low |
| Fonkeng,2015 | [53] | 2017 | 2015 | Cross-sectional | North-West and West | Unspecific TB patient | PTB | 222 | 86 (38.7) | NR | NR | NR | NR | NR | NR | Low |
| Kehbila,2015 | [54] | 2016 | 2015 | Cross-sectional | South-West | Unspecific adult TB patient | PTB | 265 | 88 (33.2) | NR | NR | NR | NR | NR | NR | Low |
|  |  |  |  |  |  |  |  |  |  | Total treated | Treatment | Success | Mortality | Failure | Loss to follow-up |  |
| Kuaban,2015 | [55] | 2020 | 2015 | Cohort | North-West, South-West, West, and Littoral | Retreated TB patient | PTB | 753 | 251 (33.3) | NR | NR | NR | NR | NR | NR | Low |
| Merker,2015 | [56] | 2021 | 2015 | Cross-sectional | North-West | RR-TB patient | PTB | 195 | 53 (27.2) | NR | NR | NR | NR | NR | NR | Low |
| Onana,2015 | [57] | 2018 | 2015 | Cross-sectional | Littoral | Unspecific TB patient | PTB | 693 | 112 (16.2) | NR | NR | NR | NR | NR | NR | Low |
| Poka-Mayap,2015 | [58] | 2020 | 2015 | Cohort | Centre | MDR-TB patient | PTB | 79 | 18 (22.8) | NR | NR | NR | NR | NR | NR | Low |
| Sama,2015 | [59] | 2017 | 2015 | Cross-sectional | North-West | Unspecific TB patient | PTB | 189 | 73 (38.6) | NR | NR | NR | NR | NR | NR | Low |
| Abanda,2016 | [60] | 2017 | 2016 | Cross-sectional | Centre, Littoral, and North-West | Unspecific adult TB patient | PTB | 270 | 63 (23.3) | NR | NR | NR | NR | NR | NR | Low |
| Bichong,2017 | [61] | 2022 | 2017 | Cross-sectional | Centre | Unspecific adult TB patient | PTB | 82 | 35 (42.7) | NR | NR | NR | NR | NR | NR | Low |
| Mbuh,2017_1 | [62] | 2021 | 2017 | Cross-sectional | Littoral | Unspecific adult TB patient | Mixed | 1665 | 440 (26.4) | NR | NR | NR | NR | NR | NR | Low |
| Mbuh,2017_2 | [63] | 2019 | 2017 | Cross-sectional | Littoral | Unspecific adult TB patient | EPTB | 45 | 13 (28.9) | NR | NR | NR | NR | NR | NR | Moderate |
| Mbuh,2017_3 | [64] | 2021 | 2017 | Cross-sectional | Littoral | RR-TB patient | PTB | 65 | 17 (26.2) | NR | NR | NR | NR | NR | NR | Low |
| Tanue,2017 | [12] | 2019 | 2017 | Cohort | Sout-West | Unspecific TB patient | NR | 2928 | 1041 (35.6) | 1041 | Standard regimen | 818 (78.6) | 72 (6.9) | 16 (1.5) | 99 (9.5) | Low |
| Nguepy,2018 | [65] | 2018 | 2018 | Cross-sectional | West | Unspecific TB patient | NR | 59 | 15 (25.4) | NR | NR | NR | NR | NR | NR | Low |
| Donfack,2019 | [66] | 2023 | 2019 | Cross-sectional | Centre and South-West | Unspecific adult TB patient | EPTB | 130 | 66 (50.8) | NR | NR | NR | NR | NR | NR | Low |
| Bitchong,2019 | [67] | 2021 | 2019 | Cross-sectional | Centre | Unspecific adult TB patient | Mixed | 75 | 48 (64.0) | NR | NR | NR | NR | NR | NR | Low |
| Jouego,2019 | [68] | 2022 | 2019 | Cohort | Multiregion | RR-TB patient | PTB | 709 | 213 (30.0) | NR | NR | NR | NR | NR | NR | Low |
| Meriki,2014 | [41] | 2019 | 2019 | Cohort | South-West | TB/HIV coinfected patients | NR | NR | NR | 209 | Standard regimen | NR | 71 (34.0) | NR | NR | Moderate |
| Mpoh,2019 | [69] | 2023 | 2019 | Cohort | Centre | MDR-TB patient | Mixed | 107 | 39 (36.4) | NR | NR | NR | NR | NR | NR | Moderate |
| Sama,2019 | [70] | 2023 | 2019 | Cross-sectional | North-West | Unspecific TB patient | PTB | 1115 | 430 (38.6) | NR | NR | NR | NR | NR | NR | Low |
|  |  |  |  |  |  |  |  |  |  | Total treated | Treatment | Success | Mortality | Failure | Loss to follow-up |  |
| Tiani,2019 | [71] | 2022 | 2019 | Cross-sectional | Centre | Unspecific adult TB patient | PTB | 336 | 99 (29.5) | NR | NR | NR | NR | NR | NR | Low |
| Djuikoue,2020 | [72] | 2022 | 2020 | Cross-sectional | Littoral | Unspecific adult TB patient | PTB | 304 | 84 (27.6) | NR | NR | NR | NR | NR | NR | Low |
| Foncha,2020 | [73] | 2021 | 2020 | Cohort | Littoral | Unspecific adult TB patient | Mixed | 3321 | 1163 (35.0) | NR | NR | NR | NR | NR | NR | Low |
| Ngangue,2020 | [74] | 2022 | 2020 | Cross-sectional | North-West, South-West, and Littoral | Unspecific adult TB patient | PTB | 251 | 74 (29.5) | NR | NR | NR | NR | NR | NR | Low |
| Chiabi,2021 | [75] | 2023 | 2021 | Cross-sectional | North-West | Pediatric | Mixed | 89 | 19 (21.3) | NR | NR | NR | NR | NR | NR | Low |
| Abessolo,2022 | [76] | 2023 | 2022 | Cross-sectional | Centre | Unspecific adult TB patient | Mixed | 692 | 416 (60.1) | 395 | Standard regimen | 311 (78.7) | 75 (19.0) | NR | NR | Low |
| Anye,2022 | [77] | 2023 | 2022 | Cross-sectional | South-West | Unspecific adult TB patient | Mixed | 375 | 76 (20.3) | NR | NR | NR | NR | NR | NR | Low |
| Chethkwo,2022 | [78] | 2024 | 2022 | Cohort | North-West | Unspecific TB patient | Mixed | 2407 | 1147 (47.7) | 1147 | Standard regimen | 963 (84.0) | 137 (11.9) | 3 (0.3) | 44 (3.8) | Low |
| Magha,2022 | [79] | 2024 | 2022 | Cross-sectional | South-West and Littoral | Unspecific adult TB patient | PTB | 549 | 69 (12.6) | NR | NR | NR | NR | NR | NR | Low |
| Mbuh,2022 | [80] | 2024 | 2022 | Cohort | Littoral | Unspecific adult TB patient | Mixed | 444 | 107 (24.1) | NR | NR | NR | NR | NR | NR | Low |
| Nchang,2022 | [81] | 2025 | 2022 | Cross-sectional | Littoral | Pediatric | PTB | 712 | 91 (12.8) | NR | NR | NR | NR | NR | NR | Low |
| Pokam,2022_1 | [82] | 2025 | 2022 | Cross-sectional | South-West | Unspecific adult TB patient | PTB | 153 | 15 (9.8) | NR | NR | NR | NR | NR | NR | Low |
| Pokam,2022_2 | [83] | 2024 | 2022 | Cross-sectional | South-West | Unspecific TB patient | Mixed | 1541 | 620 (40.2) | NR | NR | NR | NR | NR | NR | Low |
| Vukugah,2022 | [84] | 2025 | 2022 | Case-control | Centre | Pediatric | NR | 120 | 23 (19.2) | NR | NR | NR | NR | NR | NR | Low |
| Endale,2023 | [85] | 2026 | 2023 | Cross-sectional | Littoral | MDR-TB patient | PTB | 141 | 25 (17.7) | NR | NR | NR | NR | NR | NR | Low |
| Enoh,2019 | [86] | 2023 | 2023 | Cohort | South-West | Unspecific adult TB patient | NR | 74 | 20 (27.0) | NR | NR | NR | NR | NR | NR | Moderate |
| Kamdem,2023 | [87] | 2025 | 2023 | Cohort | Centre | Unspecific adult TB patient | PTB | 300 | 53 (17.7) | NR | NR | NR | NR | NR | NR | Moderate |

*1: Year of study completion; Ref: Reference; RCT: Randomized controlled trial; TB: Mycobacterium tuberculosis; RR-TB: Rifampicin-resistant Mycobacterium tuberculosis; MDR-TB: Multidrug-resistant Mycobacterium tuberculosis; NR; Not reported; PTB: Pulmonary Mycobacterium tuberculosis; EPTB: Extra-Pulmonary Mycobacterium tuberculosis*

### 3.3. HIV prevalence among tuberculosis patients

The pooled HIV prevalence rate among tuberculosis patients in Cameroon was 29.2% (95% CI: 26.3%–32.3%) based on 67 study reports including 57,169 participants. The largest study (n = 17,691) reported an HIV prevalence of 29.2% (95% CI: 28.5%–29.9%). Heterogeneity across studies was high (*I²* = 97.8%; *p* < 0.001), indicating that the pooled prevalence should be interpreted with caution. Individual study estimates ranged from 9.8% to 68.5% (**Fig. 2**). The sensitivity analysis revealed an overall robust finding with no single study significantly influencing the pooled prevalence. Moreover, despite slight asymmetry in funnel plots, no significant sign of publication bias was detected with the Egger’s test (*p* = 0.241) and Begg’s test (*p* = 0.808) (**Fig. S1-S2**).

**Fig. 2.**
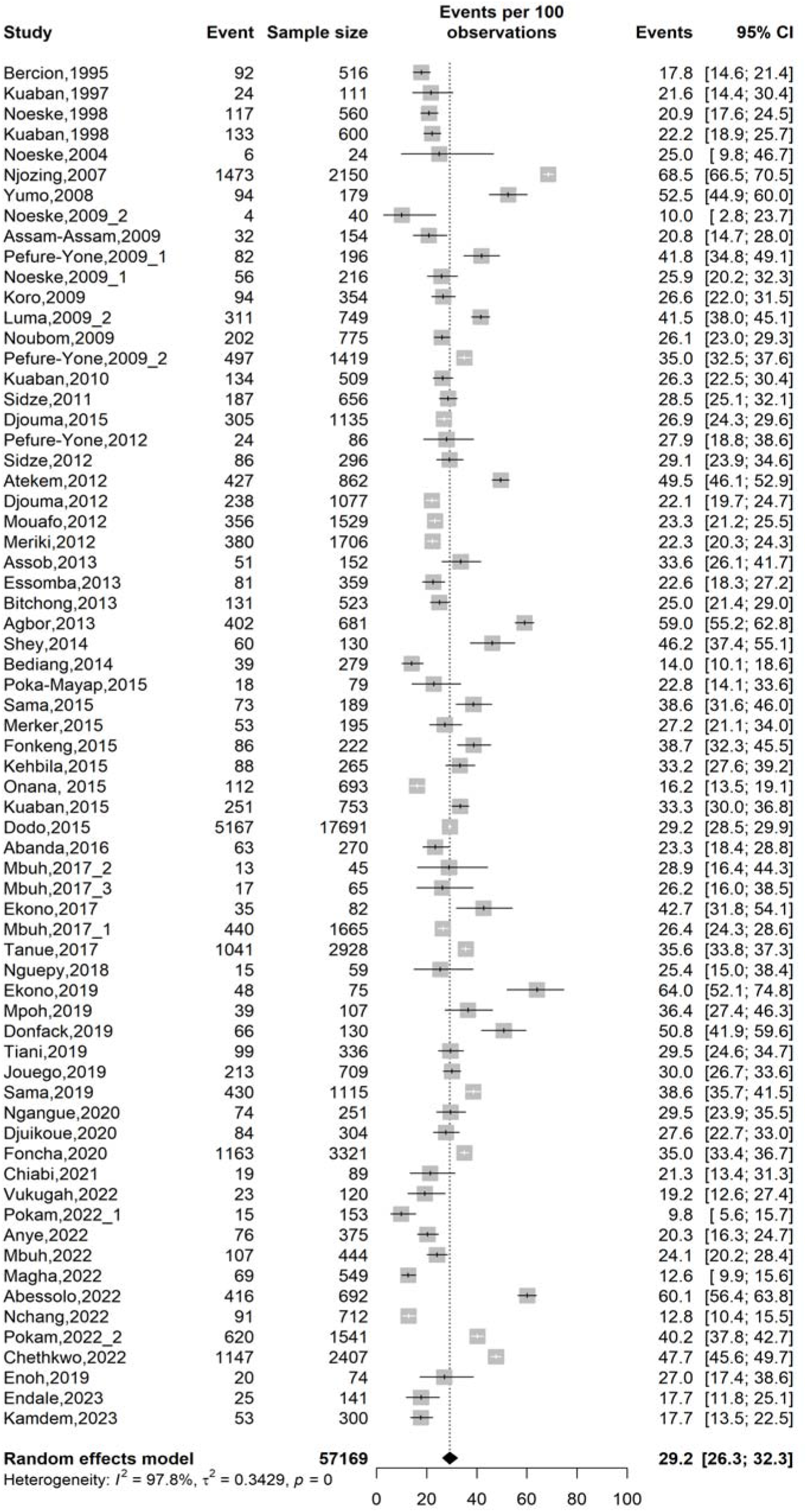
Forest plot displaying the pooled prevalence of human immunodeficiency virus infection among tuberculosis patients in Cameroon, 1995-2023

A subgroup meta-analysis examining variations in HIV prevalence among tuberculosis patients in Cameroon revealed statistically significant effect modification across all subgroups, except for risk of bias (*p* = 0.849).

Regarding study period, studies conducted before 2007 reported a significantly lower pooled HIV prevalence of 20.5% (95% CI: 18.7%–22.5%; 5 studies; n = 1,811) compared to 29.9% (95% CI: 26.8%–33.2%; 62 studies; n = 55,358) in studies from 2007 onwards (*p* < 0.001), coinciding with the WHO integration of HIV screening policies during TB clinical management.

By study design, cohort studies reported the highest prevalence at 32.5% (95% CI: 27.2%–38.3%; 20 studies; n = 36,727), followed by cross-sectional studies at 28.5% (95% CI: 25.0%–32.3%; 44 studies; n = 18,908).

Geographically, the North-West Region had the highest pooled prevalence at 41.9% (95% CI: 31.0%–53.6%; 7 studies; n = 6,324), significantly higher than the West Region (23.8%; 95% CI: 21.8%–25.9%; 6 studies; n = 4,206) and Littoral Region (25.0%; 95% CI: 21.2%–29.3%; 15 studies; n = 10,419) (*p* = 0.011) (**Fig. 3**).

**Fig. 3.**
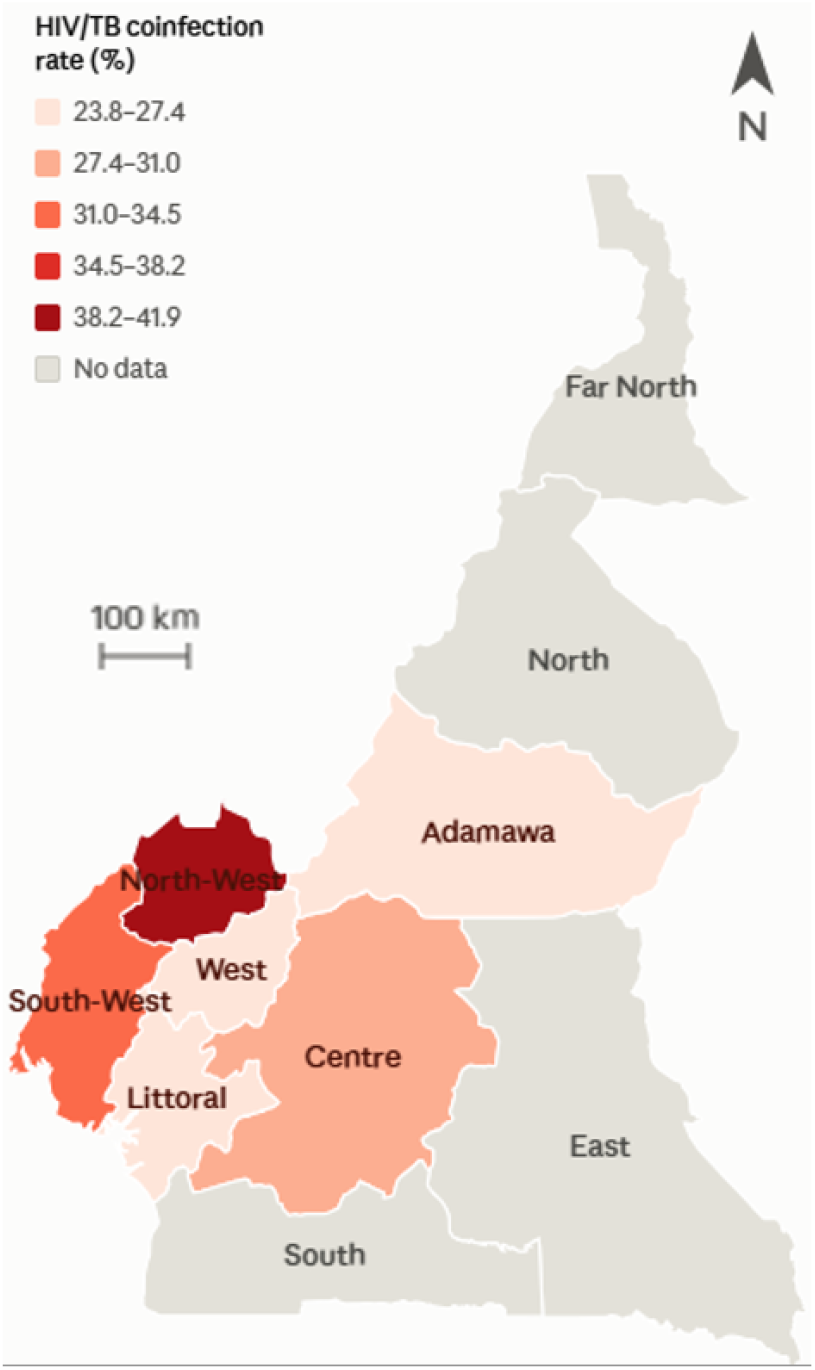
Map displaying the pooled prevalence of human immunodeficiency virus infection among tuberculosis patients in Cameroon by regions, 1995-2023

Regarding participant type, unspecific tuberculosis patients had the highest prevalence at 36.8% (95% CI: 30.4%–43.6%; 16 studies; n = 17,176), followed by RR-TB patients at 29.2% (95% CI: 26.4%–32.1%; 3 studies; n = 969), while pediatric patients had the lowest at 16.2% (95% CI: 11.9%–21.6%; 3 studies; n = 921) (*p* < 0.001).

For TB disease localization, EPTB cases had the highest HIV prevalence at 42.3% (95% CI: 33.8%–51.3%; 3 studies; n = 371), followed by mixed PTB/EPTB cases at 37.7% (95% CI: 31.5%–44.4%; 21 studies; n = 37,239), while PTB cases had the lowest at 23.9% (95% CI: 21.5%–26.6%; 37 studies; n = 16,096) (*p* < 0.001).

Heterogeneity remained high (*I*² ≥ 90%) across most subgroups, suggesting substantial variability within strata that was not explained by the subgroup variables alone, except for the pre-2007 period (*I*² = 0.0%), Adamawa region (*I*² = 0.0%), and RR-TB patients (*I*² = 0.0%), which showed consistent estimates (**Table 2** **and Figs. S3-S9**).

**Table 2.**
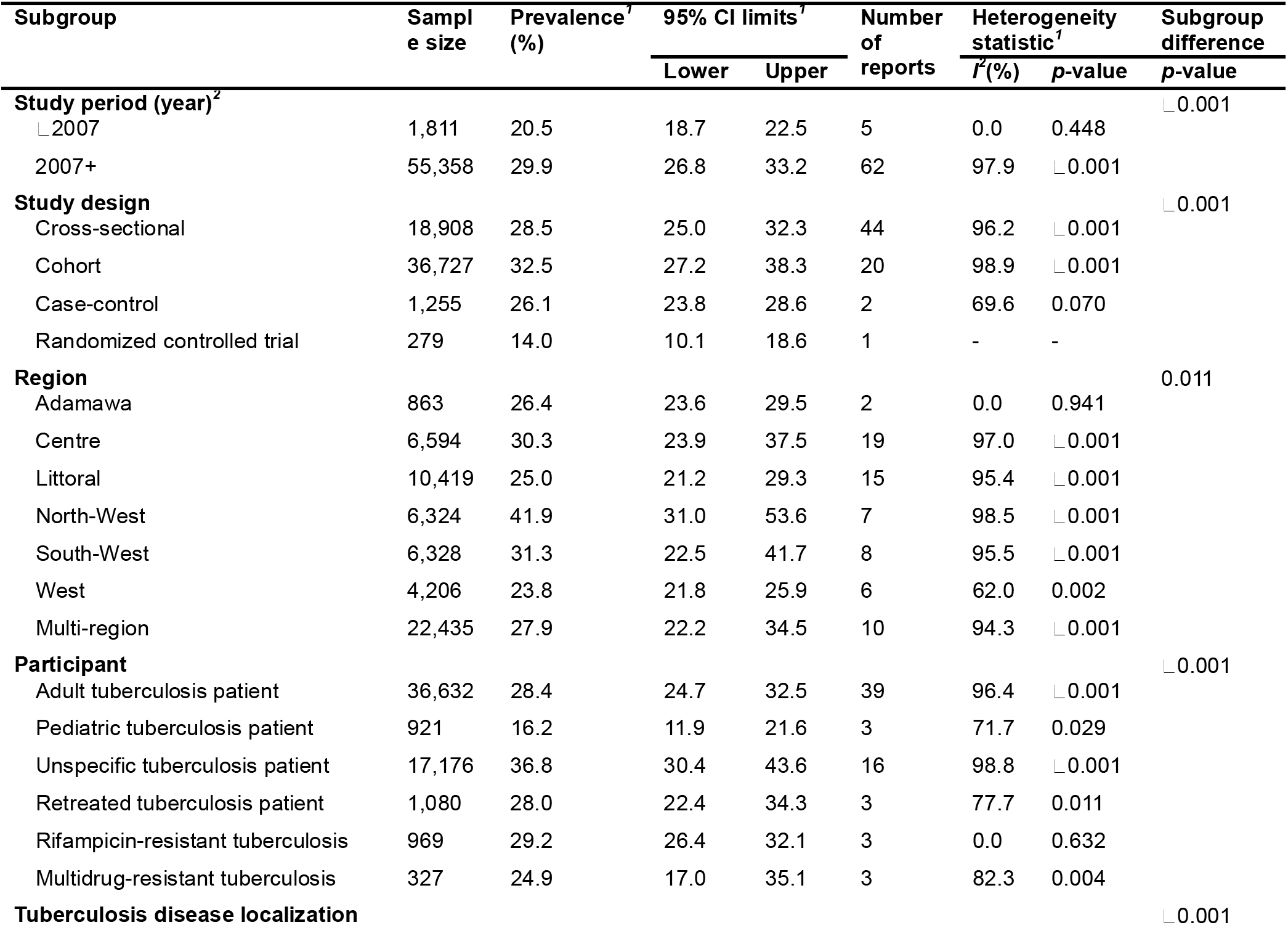
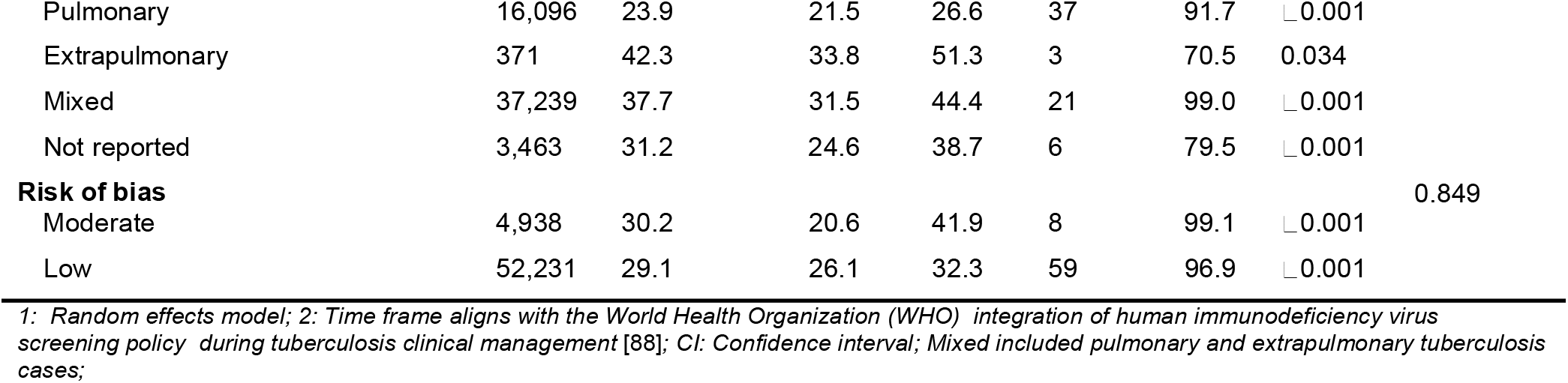
Subgroup meta-analysis of human immunodeficiency virus infection prevalence rate among tuberculosis patients in Cameroon, 1995-2023

| **Subgroup** | **Sample size** | **Prevalence<sup>*1*</sup> (%)** | **95% CI limits<sup>*1*</sup>** |  | **Number of reports** | **Heterogeneity statistic<sup>*1*</sup>** |  | **Subgroup difference** |
|---|---|---|---|---|---|---|---|---|
|  |  |  | **Lower** | **Upper** |  | ***I*<sup>*2*</sup>(%)** | ***p*-value** | ***p*-value** |
| **Study period (year)<sup>*2*</sup>** |  |  |  |  |  |  |  | ∟0.001 |
| ∟2007 | 1,811 | 20.5 | 18.7 | 22.5 | 5 | 0.0 | 0.448 |  |
| 2007+ | 55,358 | 29.9 | 26.8 | 33.2 | 62 | 97.9 | ∟0.001 |  |
| **Study design** |  |  |  |  |  |  |  | ∟0.001 |
| Cross-sectional | 18,908 | 28.5 | 25.0 | 32.3 | 44 | 96.2 | ∟0.001 |  |
| Cohort | 36,727 | 32.5 | 27.2 | 38.3 | 20 | 98.9 | ∟0.001 |  |
| Case-control | 1,255 | 26.1 | 23.8 | 28.6 | 2 | 69.6 | 0.070 |  |
| Randomized controlled trial | 279 | 14.0 | 10.1 | 18.6 | 1 | - | - |  |
| **Region** |  |  |  |  |  |  |  | 0.011 |
| Adamawa | 863 | 26.4 | 23.6 | 29.5 | 2 | 0.0 | 0.941 |  |
| Centre | 6,594 | 30.3 | 23.9 | 37.5 | 19 | 97.0 | ∟0.001 |  |
| Littoral | 10,419 | 25.0 | 21.2 | 29.3 | 15 | 95.4 | ∟0.001 |  |
| North-West | 6,324 | 41.9 | 31.0 | 53.6 | 7 | 98.5 | ∟0.001 |  |
| South-West | 6,328 | 31.3 | 22.5 | 41.7 | 8 | 95.5 | ∟0.001 |  |
| West | 4,206 | 23.8 | 21.8 | 25.9 | 6 | 62.0 | 0.002 |  |
| Multi-region | 22,435 | 27.9 | 22.2 | 34.5 | 10 | 94.3 | ∟0.001 |  |
| **Participant** |  |  |  |  |  |  |  | ∟0.001 |
| Adult tuberculosis patient | 36,632 | 28.4 | 24.7 | 32.5 | 39 | 96.4 | ∟0.001 |  |
| Pediatric tuberculosis patient | 921 | 16.2 | 11.9 | 21.6 | 3 | 71.7 | 0.029 |  |
| Unspecific tuberculosis patient | 17,176 | 36.8 | 30.4 | 43.6 | 16 | 98.8 | ∟0.001 |  |
| Retreated tuberculosis patient | 1,080 | 28.0 | 22.4 | 34.3 | 3 | 77.7 | 0.011 |  |
| Rifampicin-resistant tuberculosis | 969 | 29.2 | 26.4 | 32.1 | 3 | 0.0 | 0.632 |  |
| Multidrug-resistant tuberculosis | 327 | 24.9 | 17.0 | 35.1 | 3 | 82.3 | 0.004 |  |
| **Tuberculosis disease localization** |  |  |  |  |  |  |  | ∟0.001 |
| Pulmonary | 16,096 | 23.9 | 21.5 | 26.6 | 37 | 91.7 | ∟0.001 |  |
| Extrapulmonary | 371 | 42.3 | 33.8 | 51.3 | 3 | 70.5 | 0.034 |  |
| Mixed | 37,239 | 37.7 | 31.5 | 44.4 | 21 | 99.0 | ∟0.001 |  |
| Not reported | 3,463 | 31.2 | 24.6 | 38.7 | 6 | 79.5 | ∟0.001 |  |
| **Risk of bias** |  |  |  |  |  |  |  | 0.849 |
| Moderate | 4,938 | 30.2 | 20.6 | 41.9 | 8 | 99.1 | ∟0.001 |  |
| Low | 52,231 | 29.1 | 26.1 | 32.3 | 59 | 96.9 | ∟0.001 |  |

*1: Random effects model; 2: Time frame aligns with the World Health Organization (WHO) integration of human immunodeficiency virus screening policy during tuberculosis clinical management* [88]*; CI: Confidence interval; Mixed included pulmonary and extrapulmonary tuberculosis cases;*

### 3.4. Treatment outcome among HIV/tuberculosis coinfected patients

Among HIV/TB co-infected patients, the pooled treatment success rate was 75.9% (95% CI: 70.4–80.8; 8 studies; n = 4,230), while the pooled mortality rate was 21.1% (95% CI: 11.9– 34.7; 10 studies; n = 4,403). Treatment failure was low, with a pooled estimate of 0.3% (95% CI: 0.1–1.5; 6 studies; n = 3,520), and the pooled loss to follow-up rate was 7.7% (95% CI: 4.3–13.3; 5 studies; n = 3,103). However, all pooled estimates demonstrated substantial heterogeneity across studies, warranting cautious interpretation of these findings (**Figs. 4-7**).

**Fig. 4.**
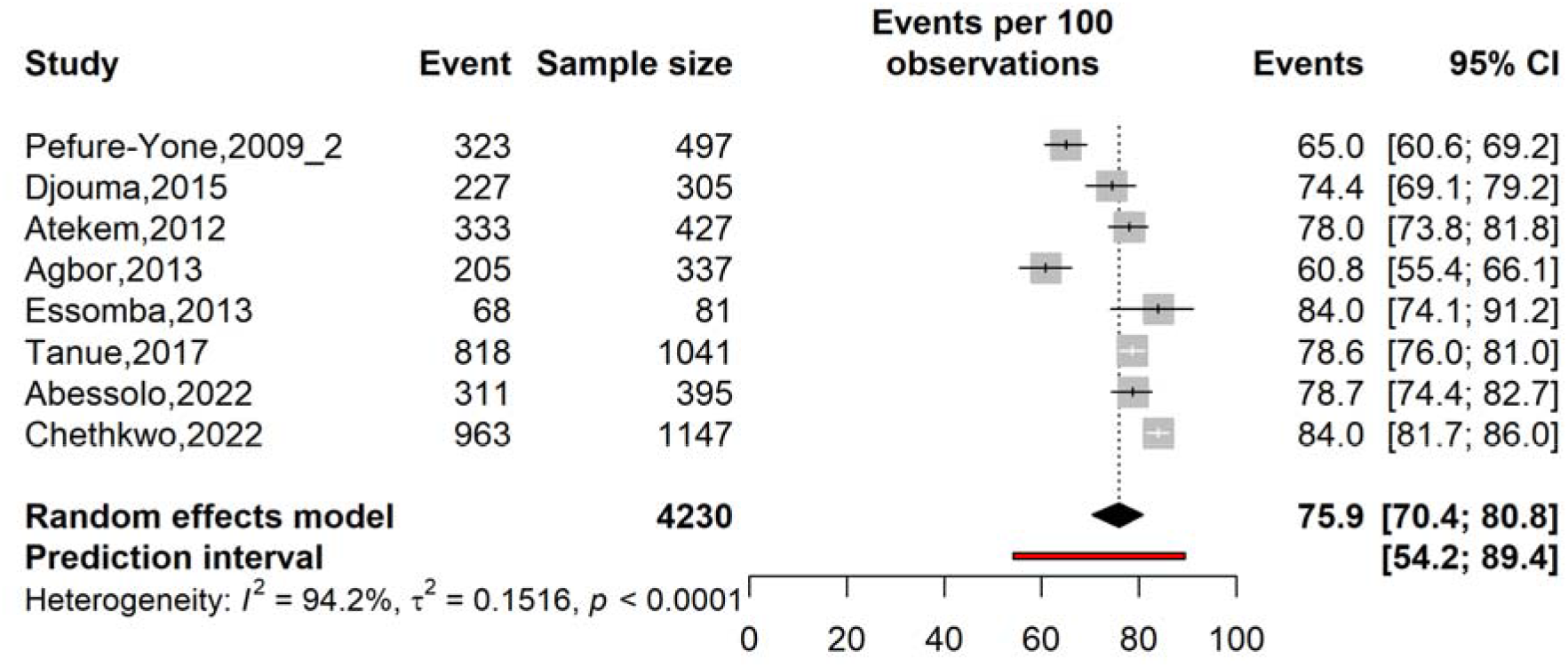
Forest plot displaying the pooled tuberculosis treatment success rate among patients coinfected with human immunodeficiency virus in Cameroon, 2009-2022

**Fig. 5.**
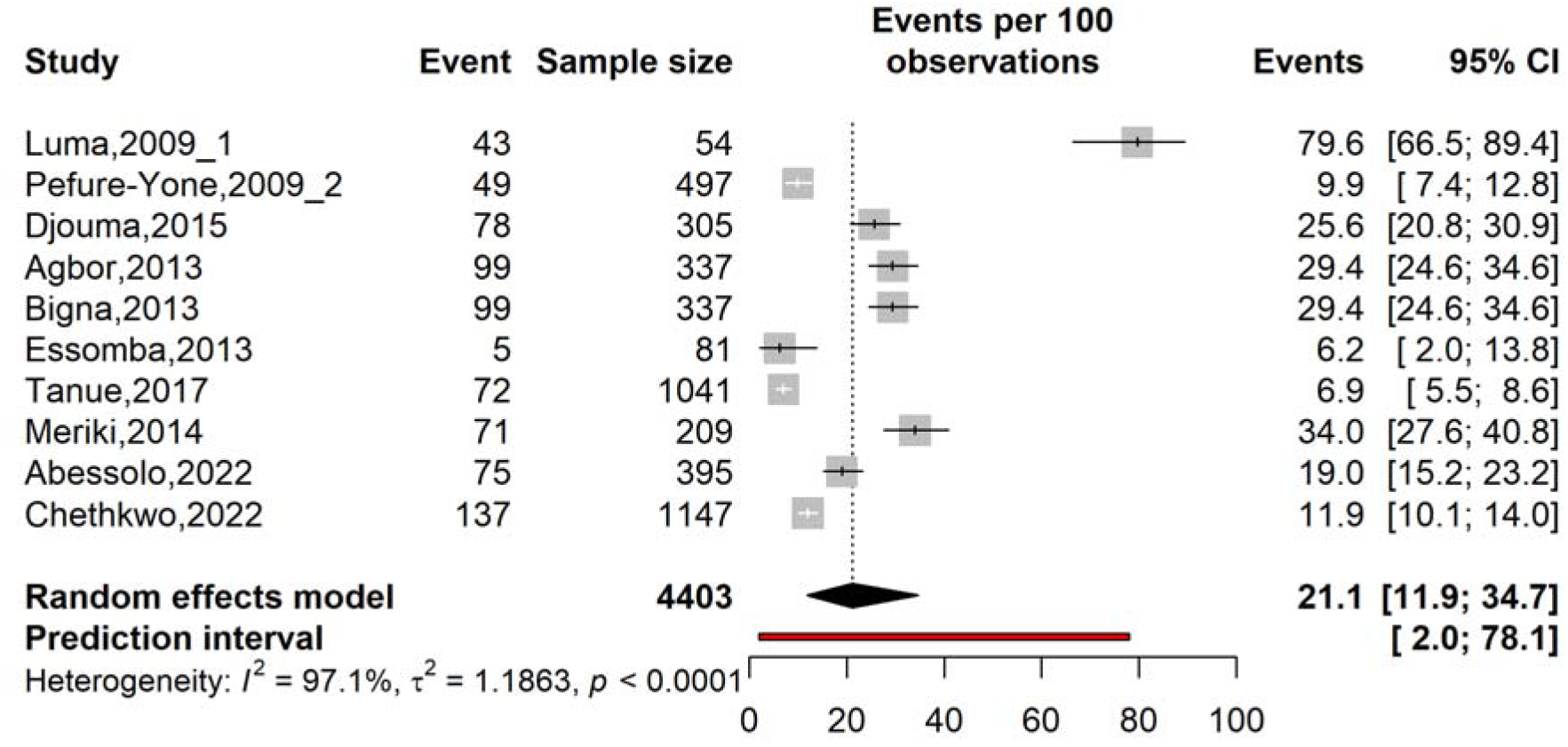
Forest plot displaying the pooled tuberculosis mortality rate among patients coinfected with human immunodeficiency virus in Cameroon, 2009-2022

**Fig. 6.**
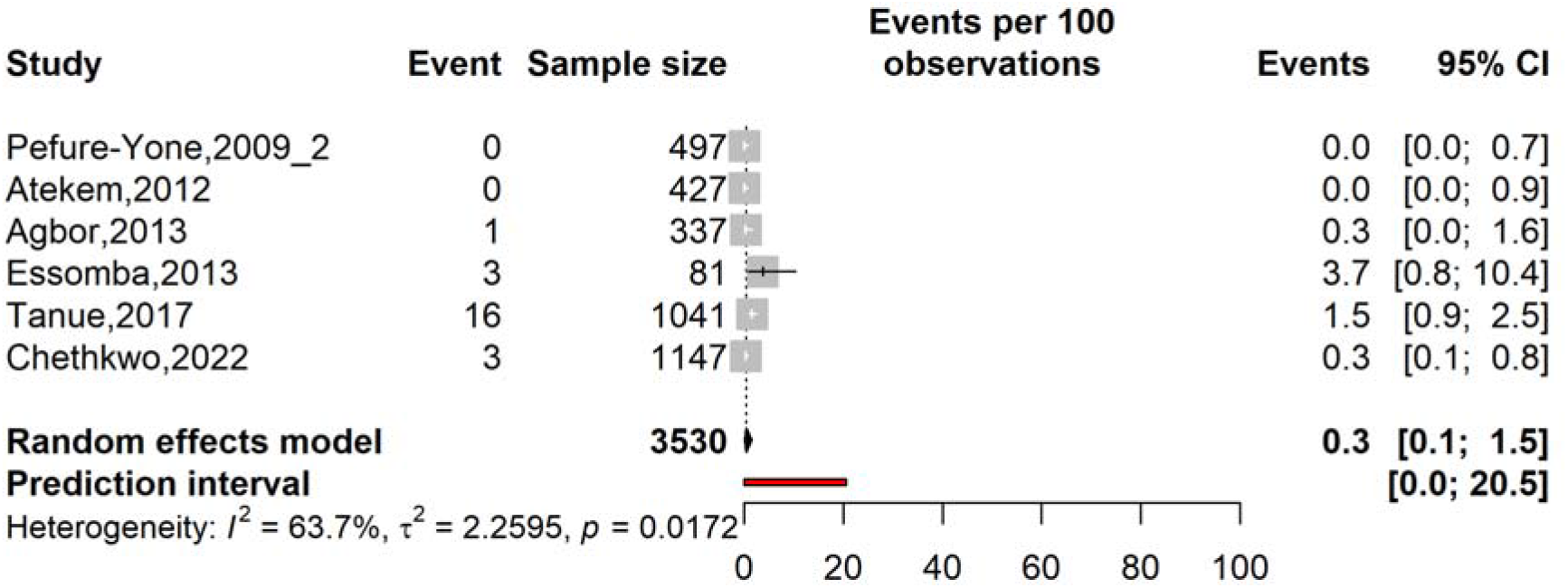
Forest plot displaying the pooled tuberculosis treatment failure rate among patients coinfected with human immunodeficiency virus in Cameroon, 2009-2022

**Fig. 7.**
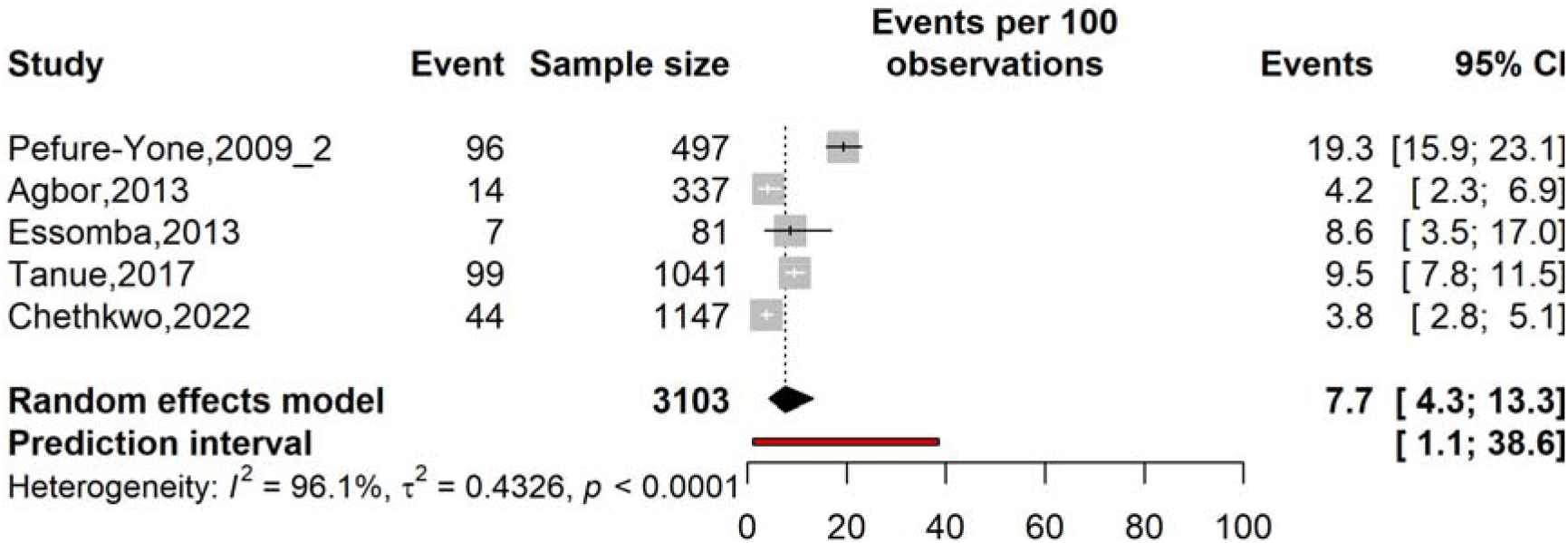
Forest plot displaying the pooled tuberculosis treatment loss to follow-up rate among patients coinfected with human immunodeficiency virus in Cameroon, 2009-2022

### 3.5. Predictors of unfavorable treatment outcomes

Pooled analysis of adjusted odds ratios from two studies identified the absence of antiretroviral therapy as a significant predictor of unfavorable outcomes among HIV/TB co- infected patients. HIV/TB co-infected patients not receiving antiretroviral therapy had 90% higher odds of unfavorable outcomes compared to those on treatment (AOR = 1.90; 95% CI: 1.15–3.12) (**Table 3**).

**Table 3.**
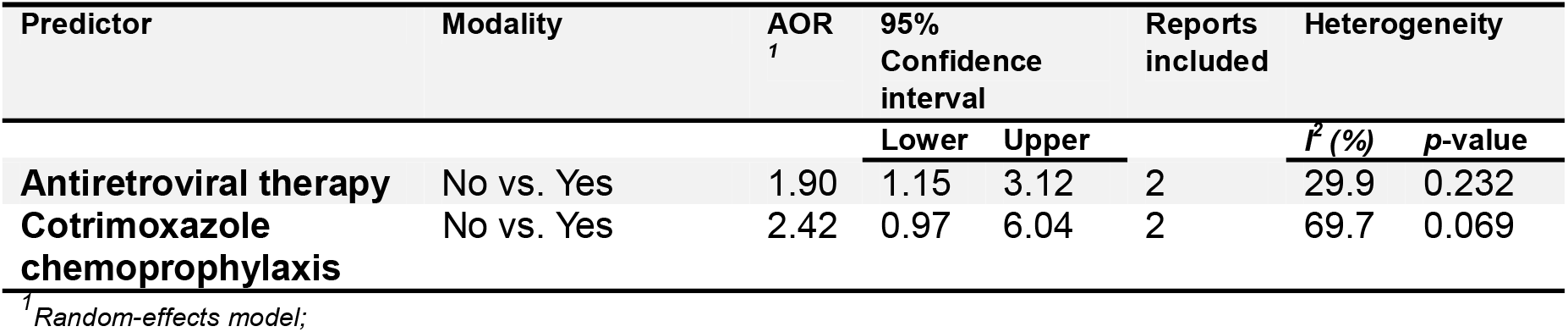
Predictors of unfavorable outcome among human immunodeficiency virus and tuberculosis coinfected patients in Cameroon, 2013-2017

## **4.** Discussion

This systematic review and meta-analysis illustrate that HIV remains a major comorbidity among people with tuberculosis in Cameroon. Overall, 29.2% of TB patients were living with HIV, meaning that almost one in three patients in the available studies had both infections. Although there was considerable variation between studies, with prevalence estimates ranging from 9.8% to 68.5%, the pooled estimate remained stable during sensitivity analysis, and there was no evidence of significant publication bias. The prevalence found in Cameroon is close to the 31.8% reported in an earlier meta-analysis of TB patients across sub-Saharan Africa [8], but higher than estimates from Nigeria (25.8%) and Ethiopia (22.0%) [89, 90]. A recent global meta-analysis reported an overall TB/HIV coinfection prevalence of 14%, with an estimate of 22% for Africa. However, that review included studies examining coinfection from both the TB and HIV populations, making direct comparison with our estimate among TB patients difficult [5]. Taken together, these findings confirm that HIV/TB coinfection continues to represent an important public health problem in Cameroon.

We also found a clear difference according to study period. HIV prevalence among TB patients was 20.5% in studies conducted before 2007, compared with 29.9% in studies conducted from 2007 onward. This should not necessarily be interpreted as a true increase in HIV/TB coinfection over time. An important change around this period was the wider integration of HIV testing into routine clinical care. In 2007, WHO and UNAIDS issued guidance promoting provider-initiated HIV testing, including testing in tuberculosis services, and HIV testing among TB patients subsequently expanded considerably across Africa [91]. Greater testing may therefore have resulted in more HIV-positive TB patients being identified in later studies. The very high heterogeneity among studies conducted after 2007 also suggests that these studies captured a wider range of populations and clinical settings.

Marked geographical differences were also observed within Cameroon. The North-West Region had the highest pooled prevalence at 41.9%, followed by the South-West and Centre Regions, while the West and Littoral Regions had lower estimates. Regional variation has also been reported elsewhere in Africa. In Nigeria, for example, pooled HIV/TB prevalence ranged from 19.3% in the South-East to 34.3% in the North-Central region [89]. Several factors may contribute to these differences, including the underlying HIV burden, access to HIV and TB services, referral patterns, and differences in the populations included in individual studies. However, heterogeneity remained high across most regional subgroups in our analysis, so these pooled estimates should be interpreted with caution and not treated as precise estimates of the current prevalence in every region.

Differences were also observed across the populations studied. Pediatric TB patients had a pooled HIV prevalence of 16.2%, compared with 28.4% among adult TB patients, while estimates were higher in some other patient categories. The lower prevalence among children should be interpreted with caution because only three pediatric studies were available. Differences between patient groups may reflect variation in exposure to HIV, routes of HIV acquisition, clinical characteristics, and the types of facilities from which participants were recruited. The small number of studies in several subgroups also limits the strength of conclusions that can be drawn from these comparisons.

Another important finding was the association between HIV and the clinical presentation of tuberculosis. HIV prevalence was 42.3% among patients with extrapulmonary TB, compared with 37.7% in studies including both pulmonary and extrapulmonary disease and 23.9% among patients with pulmonary TB alone. This pattern is consistent with the known effect of HIV-related immunosuppression on TB presentation. People living with HIV, particularly those with advanced disease, are more likely to develop extrapulmonary or disseminated tuberculosis [92]. Clinically, this reinforces the need for a high index of suspicion for extrapulmonary TB among people living with HIV, especially where conventional pulmonary investigations are inconclusive.

Treatment outcomes among coinfected patients were also concerning. The pooled treatment success rate was 75.9%, while mortality was 21.1%, loss to follow-up was 7.7%, and treatment failure was relatively low at 0.3%. The treatment success estimate is slightly below the 79% pooled treatment success rate reported in a systematic review of TB treatment outcomes across Africa. That review also found HIV coinfection to be associated with a higher risk of unsuccessful treatment [93]. Moreover, this estimate was lower than the 2022 treatment success rate objective (88%) of the National Strategic Plan against Tuberculosis in Cameroon (NSPTC) for all TB cases [94]. The mortality observed in our analysis is substantial and is comparable with findings from Ethiopia, where recent meta- analyses reported mortality of approximately 18–19% among HIV/TB-coinfected patients [95]. Differences between studies may reflect variation in the severity of HIV disease, timing of TB diagnosis, access to ART, adherence, TB presentation, and health-system factors. However, this mortality rate was lower than the 2022 objectives of the 2024–2026 NSPTC (28%) for all TB cases [94]. This finding might suggest that current strategies to reduce mortality among TB patients are yielding favorable results.

The association with antiretroviral therapy was particularly important. Patients who were not receiving ART had 90% higher odds of an unfavorable treatment outcome compared with those receiving ART (AOR = 1.90; 95% CI: 1.15–3.12). This finding supports the importance of integrating HIV and TB care and ensuring timely initiation and continuation of ART. Evidence from other African settings has similarly shown poorer outcomes among coinfected patients with inadequate ART adherence, while poor ART adherence has also been associated with increased mortality[96]. However, our pooled estimate was based on only two studies, and the magnitude of the association should therefore be interpreted cautiously. Even so, the direction of the finding is consistent with the established role of ART in improving outcomes among people with HIV-associated TB.

The high level of heterogeneity across the analysis deserves particular attention. HIV prevalence varied widely between individual studies, and heterogeneity remained above 90% in most subgroup analyses. Similar levels of heterogeneity have been reported in meta- analyses from sub-Saharan Africa, Nigeria, and Ethiopia [5, 89, 95, 97]. This probably reflects genuine differences in study populations, geographical settings, HIV epidemiology, TB presentation, diagnostic approaches, study periods, and access to health services. Importantly, prevalence did not differ significantly across risk-of-bias categories in our subgroup analysis, suggesting that the observed variability cannot be explained solely by differences in study quality. The overall estimate of 29.2% should therefore be viewed as a national summary of a highly diverse body of evidence rather than a prevalence that applies uniformly throughout Cameroon.

These findings have practical implications for TB and HIV services in Cameroon. Routine HIV testing among people with presumptive or confirmed TB remains important, particularly in settings where coinfection is common. The high prevalence among patients with extrapulmonary TB also supports careful assessment for atypical and disseminated disease among people living with HIV. Equally important is ensuring that patients diagnosed with both infections are linked promptly to ART and supported throughout TB and HIV treatment. The regional differences observed in this review suggest that TB/HIV interventions may also need to respond to local patterns rather than relying entirely on a uniform national approach.

## **5.** Limitations

This review has several limitations. Most included studies were observational, and substantial heterogeneity persisted even after subgroup analysis. Some regions and patient groups were represented by relatively few studies, limiting the precision of those subgroup estimates. The long study period also encompassed major changes in HIV testing, ART availability, TB diagnostics, and national treatment policies, making direct comparisons between older and more recent studies difficult. Treatment outcomes were reported by considerably fewer studies than HIV prevalence, and the pooled analysis of ART and unfavorable outcomes was based on only two studies. In addition, several potentially important patient-level factors could not be examined consistently because they were not reported in enough studies. The included reports were unevenly distributed across the ten regions of Cameroon, with four regions (Far-North, East, and South) not represented. These limitations should be considered when interpreting the pooled estimates and their applicability to the current epidemiology of HIV/TB coinfection in Cameroon.

## **6.** Conclusions

HIV/TB coinfection remains a significant public health concern in Cameroon, with available evidence indicating that approximately three in ten TB patients are coinfected with HIV. The burden varied considerably across regions and clinical groups and was particularly high among patients with extrapulmonary TB. Treatment outcomes remain a concern, particularly the substantial mortality observed among coinfected patients. However, the observed mortality rate aligns with the objectives of the Cameroon National Strategic Plan, suggesting that current case management strategies are encouraging. The absence of ART was associated with poorer outcomes. These findings reinforce the need for continued integration of TB and HIV services, routine HIV testing among TB patients, timely initiation and continuation of ART, and sustained follow-up throughout treatment. Further studies using more recent and geographically representative data would also help provide a clearer picture of the current burden of HIV/TB coinfection across Cameroon.

## Abbreviations

AIDS: Acquired immunodeficiency syndrome
AJOL: African Journals Online
AOR: Adjusted odds ratio
ART: Antiretroviral therapy
CD4: CD4 T-lymphocyte (cell count)
CI: Confidence interval
CPT: Cotrimoxazole preventive therapy
EPTB: Extrapulmonary tuberculosis
GLMM: Generalized linear mixed models
HIV: Human immunodeficiency virus
*I*²: Higgins’ heterogeneity statistic
JBI: Joanna Briggs Institute
LTFU: Lost to follow-up
MeSH: Medical Subject Headings
NSPTC: National Strategic Plan against Tuberculosis in Cameroon
PCR: Polymerase chain reaction
PLOGIT: Probit-logit transformation
PRISMA: Preferred Reporting Items for Systematic Reviews and Meta-Analyses
PTB: Pulmonary tuberculosis
RR-TB: Rifampicin-resistant tuberculosis
RCT: Randomized controlled trial
TB: Tuberculosis
UNAIDS: Joint United Nations Programme on HIV/AIDS
WHO: World Health Organization

## Supplementary Information

Supplementary Material

## Data Availability

All data generated in this study are available in the main manuscript and its supplementary material.

## Acknowledgment

Not applicable.

## Authors’ contributions

FZLC conceived the original idea of the study; FZLC and RT conducted the literature search; FZLC, RT, ADT, and CA selected the studies, extracted the relevant information, critically assessed included reports and synthesized the data; FZLC performed the analyses; FZLC, RT, ADT, and CA wrote the first draft of the manuscript. FZLC, RT, ADT, CA, CJKM, GCMF, AN, and SM critically reviewed and revised successive drafts of the manuscript. FZLC, ADT, CA, RT, CJKM, GCMF, AN, and SM, and MNT read and approved the final version of the manuscript.

## Declarations

Ethical approval and consent to participate

Not applicable.

## Consent for publication

Not applicable.

## Clinical trial number

Not applicable.

