## Supplementary Material for "Human immunodeficiency virus and tuberculosis coinfection prevalence and treatment outcomes in Cameroon: a systematic review and meta-analysis"

**Tables S1** Search strategy by databases

| **Database** | **Search string** | **Number of entries** |
| --- | --- | --- |
| **Pubmed** | ("HIV"[Mesh] OR "HIV"[tiab] OR "Human Immunodeficiency Virus"[tiab] OR "AIDS"[Mesh] OR "AIDS"[tiab]) AND ("Tuberculosis"[Mesh] OR "Tuberculosis"[tiab] OR "TB"[tiab] OR "Mycobacterium tuberculosis"[tiab]) AND ("Cameroon"[Mesh] OR "Cameroon"[tiab] OR "Cameroun"[tiab] OR "Cameroonian"[tiab]) | 182 |
| **Scopus** | TITLE-ABS-KEY("HIV" OR "Human Immunodeficiency Virus") AND TITLE-ABS-KEY("Tuberculosis" OR "TB" OR "Mycobacterium tuberculosis") AND TITLE-ABS-KEY("Cameroon" OR "Cameroun" OR "Cameroonian") | 239 |
| **Web of sciences** | (((((TS = HIV) OR (TS = Human Immunodeficiency Virus)) OR (TS = AIDS)) AND (((TS = Tuberculosis) OR (TS = TB)) OR (TS = Mycobacterium tuberculosis))) AND (((TS = Cameroon) OR (TS = Cameroun)) OR (TS = Cameroonian))) | 276 |
| **Embase** | ('human immunodeficiency virus infection'/exp OR  'acquired immune deficiency syndrome'/exp OR  HIV:ti,ab,kw OR 'Human Immunodeficiency Virus':ti,ab,kw OR  AIDS:ti,ab,kw) AND ('tuberculosis'/exp OR 'Mycobacterium tuberculosis'/exp OR Tuberculosis:ti,ab,kw OR  TB:ti,ab,kw OR 'Mycobacterium tuberculosis':ti,ab,kw)  AND ('cameroon'/exp OR Cameroon:ti,ab,kw OR Cameroun:ti,ab,kw OR Cameroonian*:ti,ab,kw) | 521 |
| **Cochrane Library** | (HIV OR Human Immunodeficiency Virus OR AIDS) AND (Tuberculosis OR TB OR Mycobacterium tuberculosis) AND (Cameroon OR Cameroun OR Cameroonian) | 17 |
| **AJOL** | (HIV OR Human Immunodeficiency Virus OR AIDS) AND (Tuberculosis OR TB OR Mycobacterium tuberculosis) AND (Cameroon OR Cameroun OR Cameroonian) | 18 |

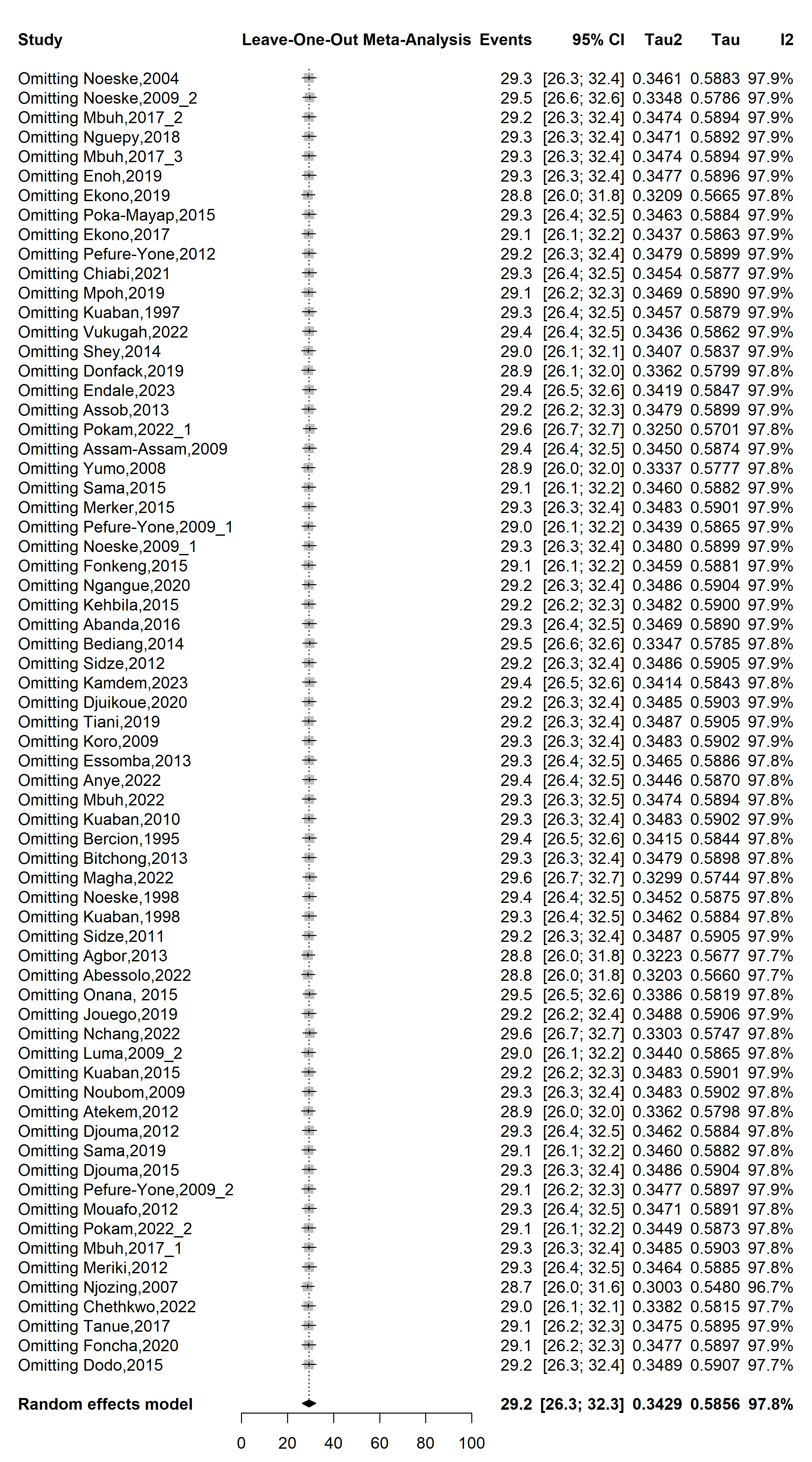

**Fig. S1** Forest plot displaying the sensitivity analysis of the pooled HIV prevalence among tuberculosis patients in Cameroon, 1995-2023

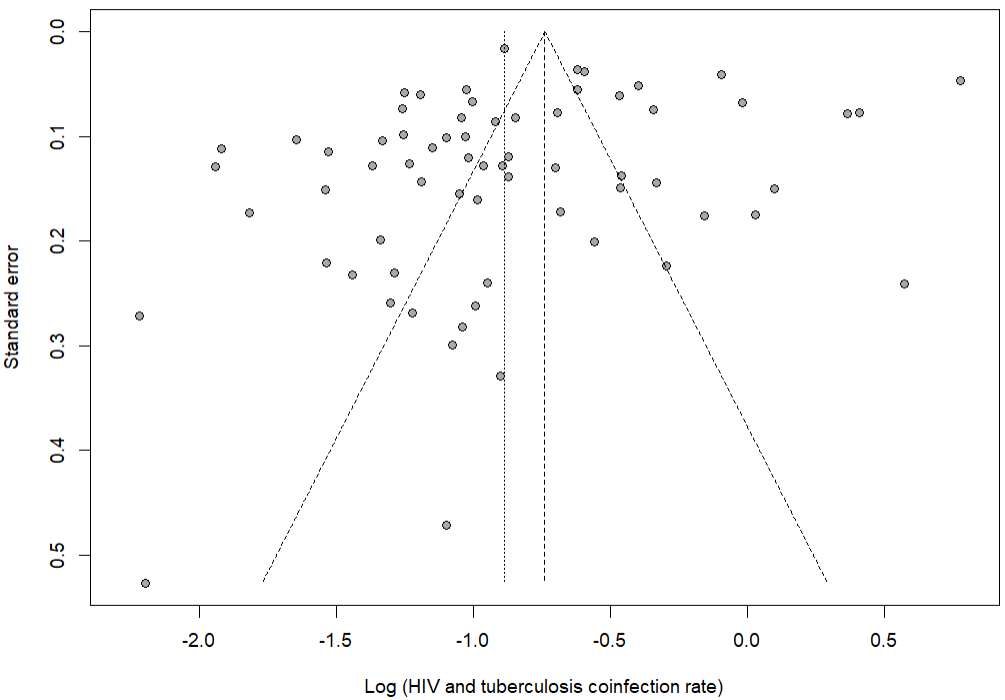

Egger’s test *p*-value = 0.241

Begg’s test *p*-value = 0.808

**Fig. S2** Funnel plot assessing publication bias among studies reporting HIV prevalence among tuberculosis patients in Cameroon, 1995-2023

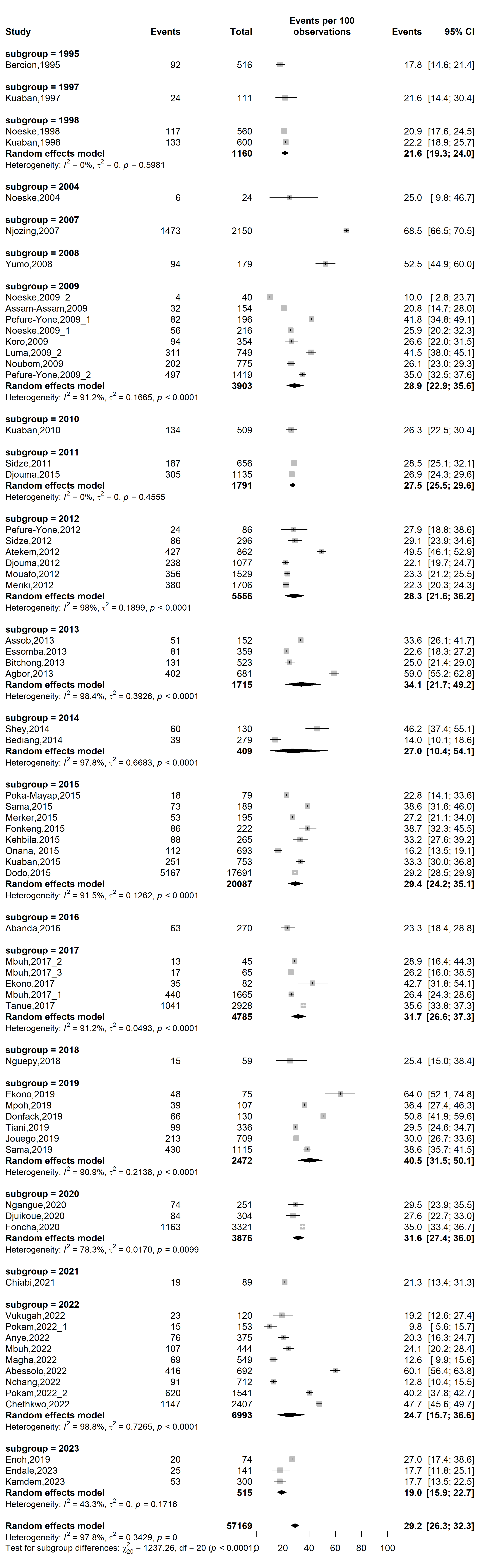

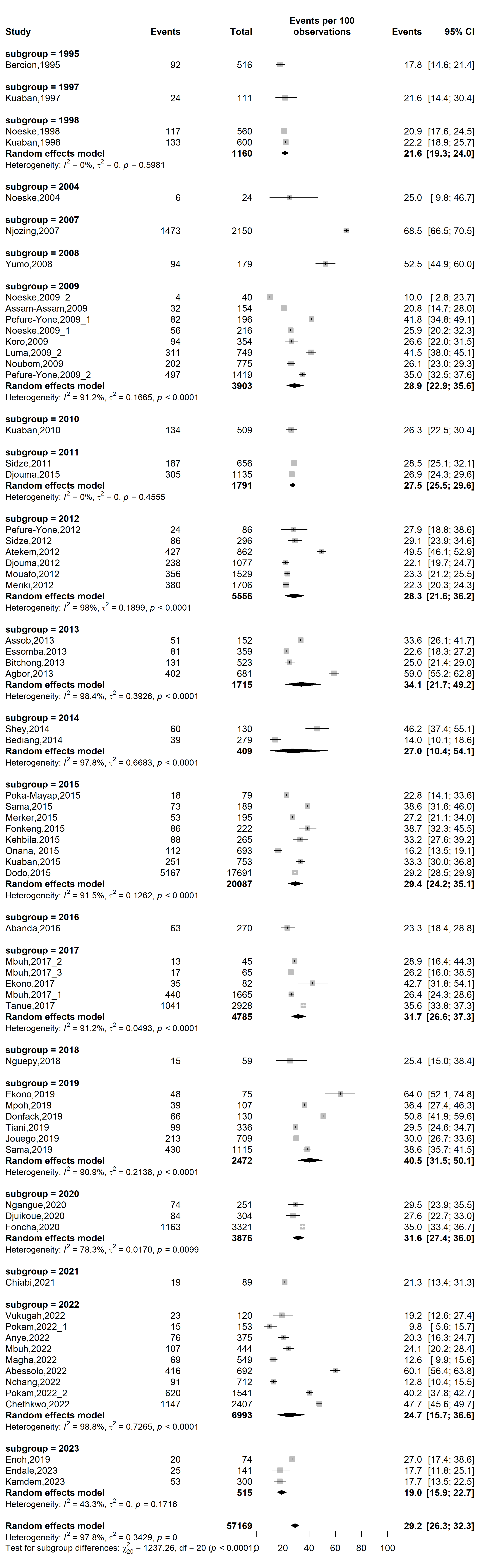

**Fig. S3** Forest plot displaying the subgroup meta-analysis of the pooled HIV prevalence among drug-resistant tuberculosis patients in Cameroon by study timeframe, 1995-2023

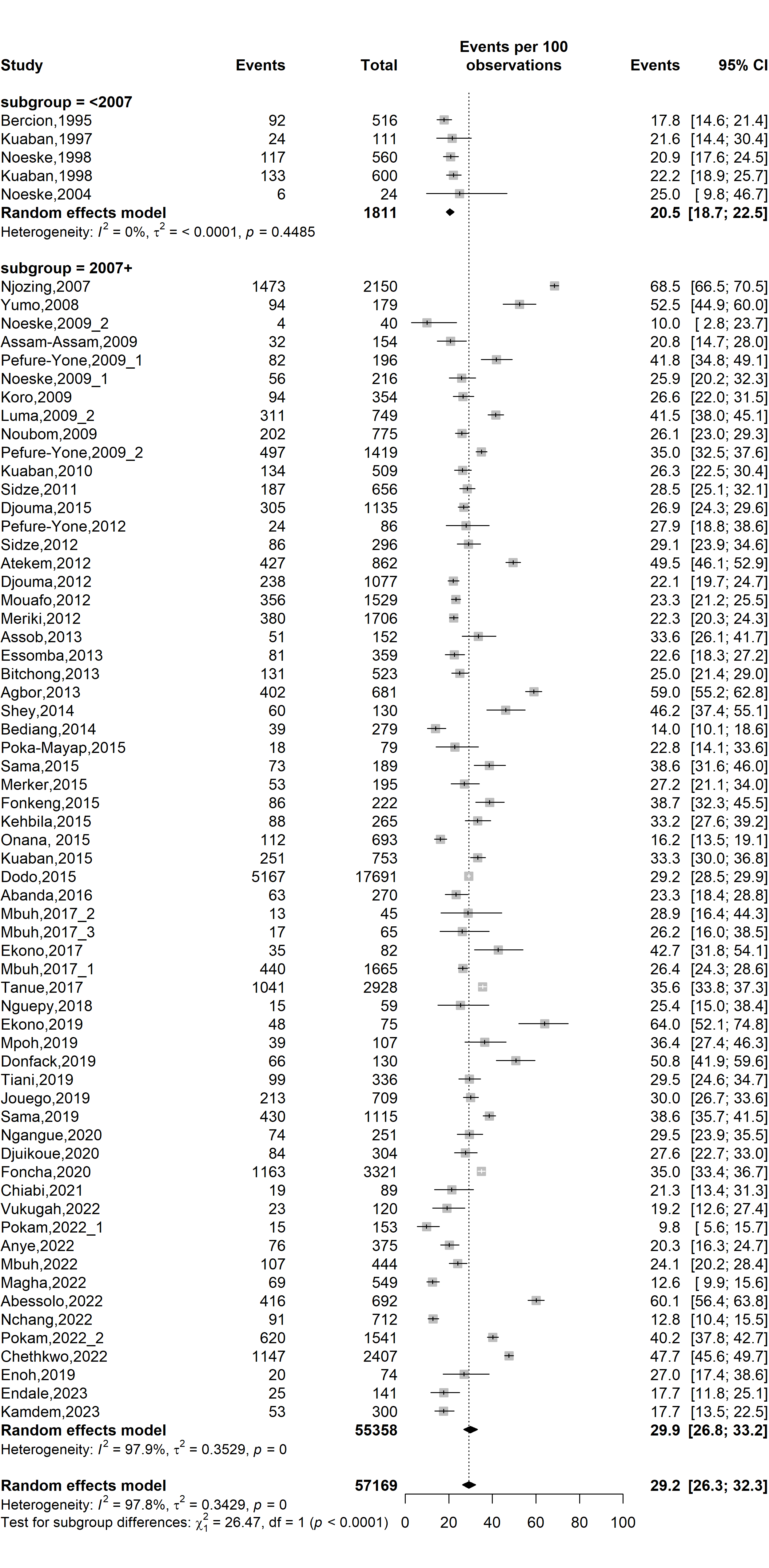

**Fig. S4** Forest plot displaying the subgroup meta-analysis of pooled prevalence of HIV prevalence among tuberculosis patients in Cameroon by HIV testing strategy, 1995-2023

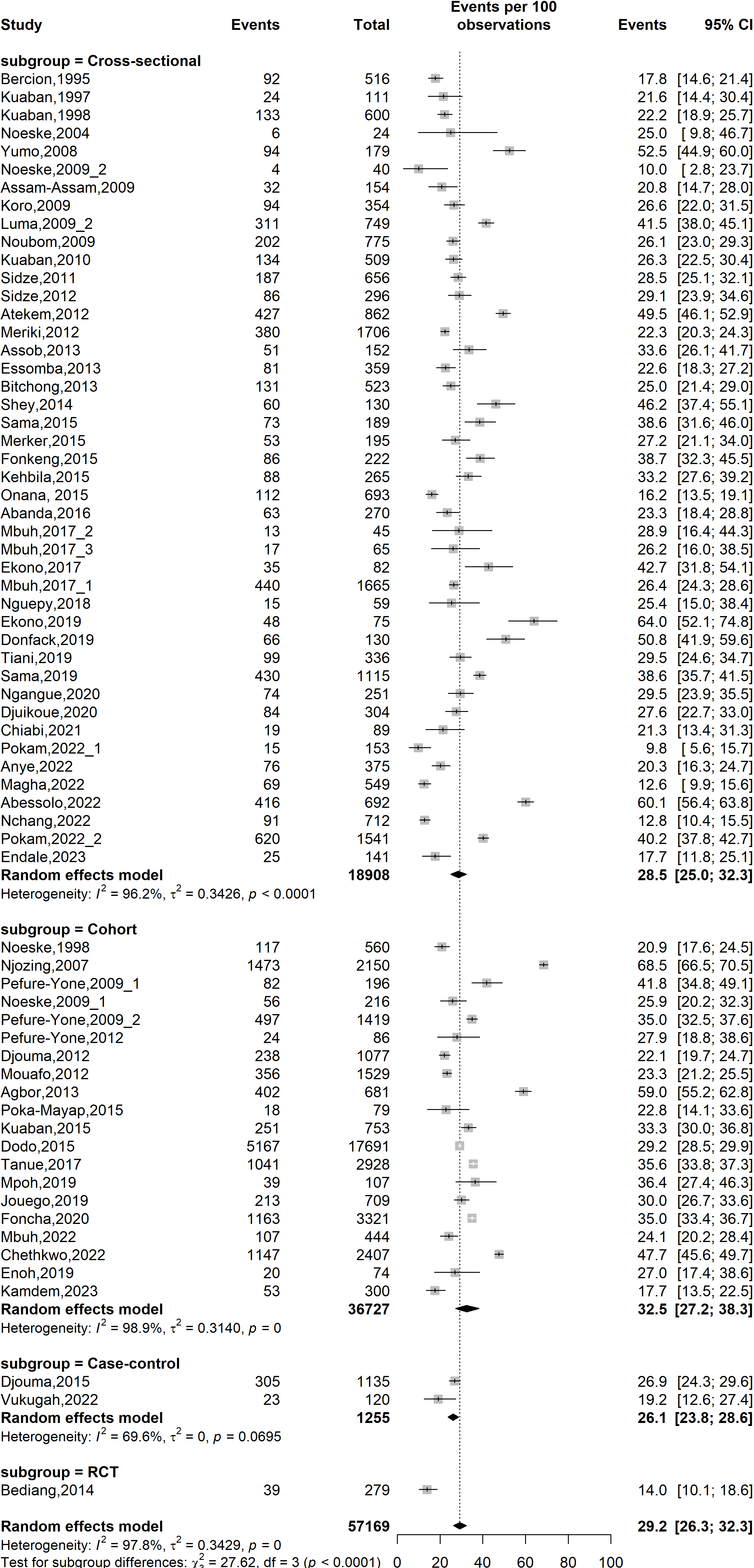

**Fig. S5** Forest plot displaying the subgroup meta-analysis of pooled prevalence of HIV prevalence among tuberculosis patients in Cameroon by study designs, 1995-2023

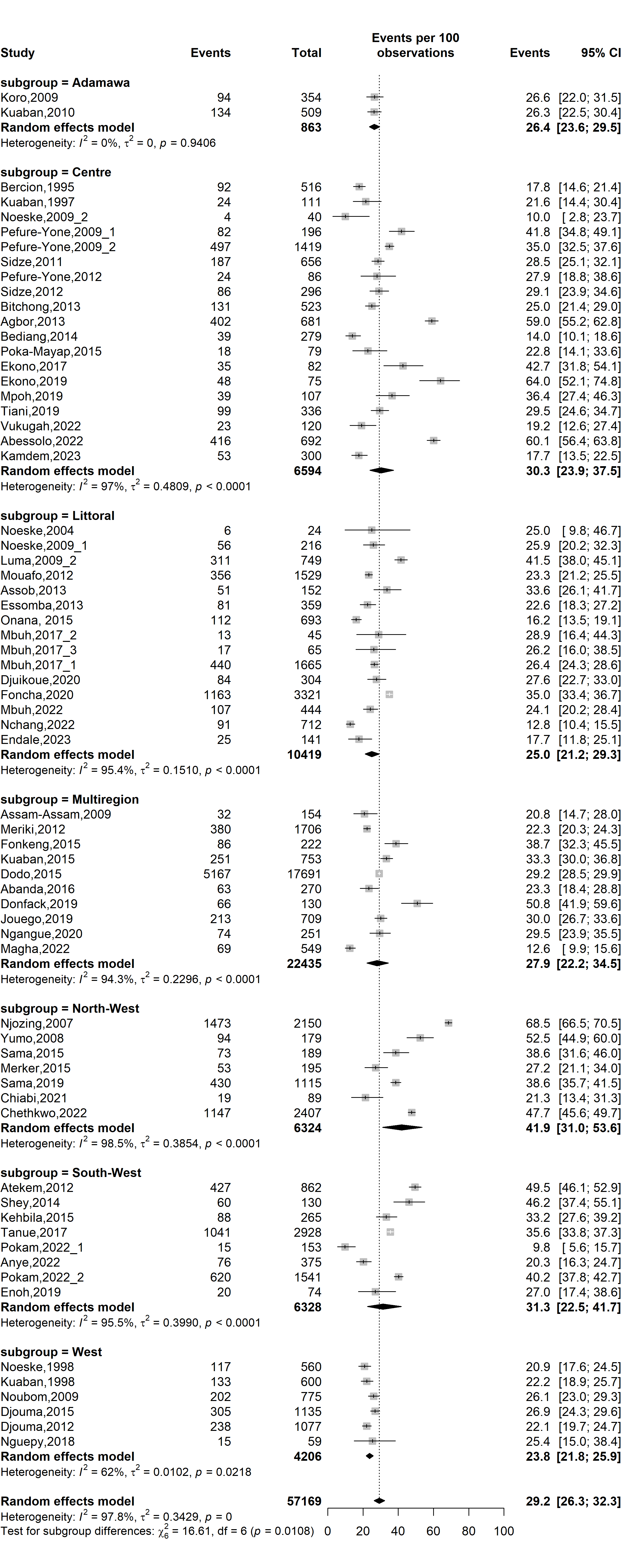

**Fig. S6** Forest plot displaying the subgroup meta-analysis of pooled prevalence of HIV prevalence among tuberculosis patients in Cameroon by study regions, 1995-2023

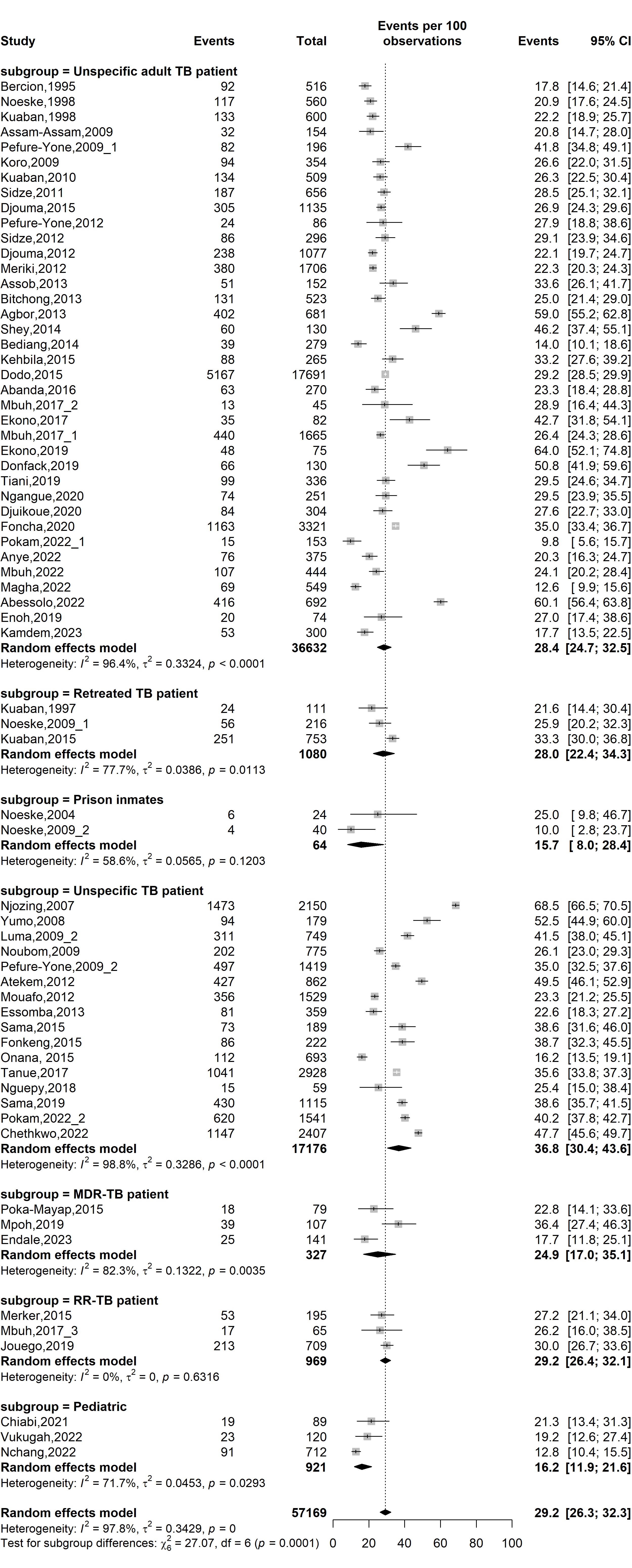

**Fig. S7** Forest plot displaying the subgroup meta-analysis of pooled prevalence of HIV prevalence among tuberculosis patients in Cameroon by types of patients, 1995-2023

*(TB: Mycobacterium tuberculosis infection; RR-TB: Rifampicin-resistant tuberculosis; MDR-TB: Multidrug-resistant tuberculosis)*

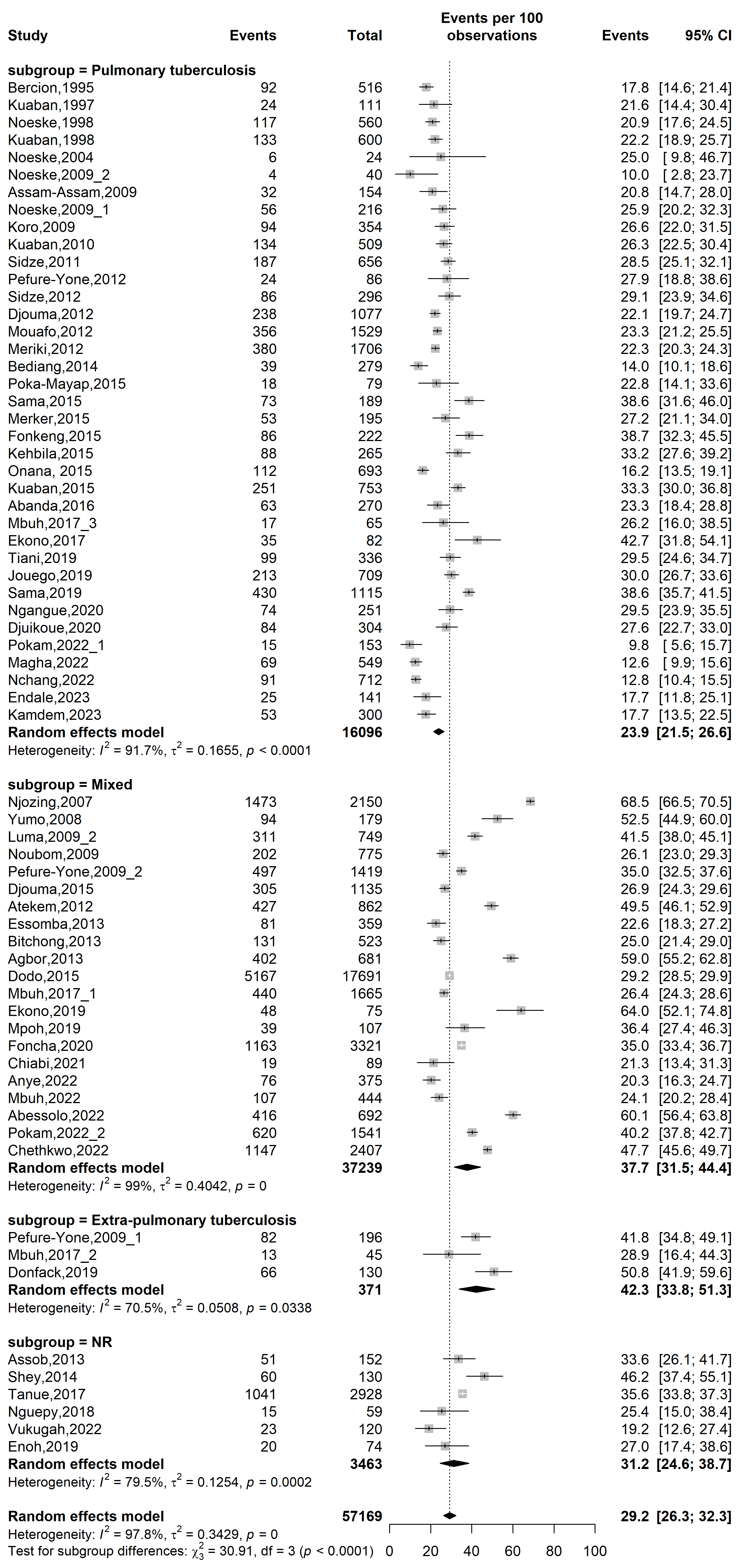

**Fig. S8** Forest plot displaying the subgroup meta-analysis of pooled prevalence of HIV prevalence among tuberculosis patients in Cameroon by tuberculosis disease localizations, 1995-2023

(NR: Not reported)

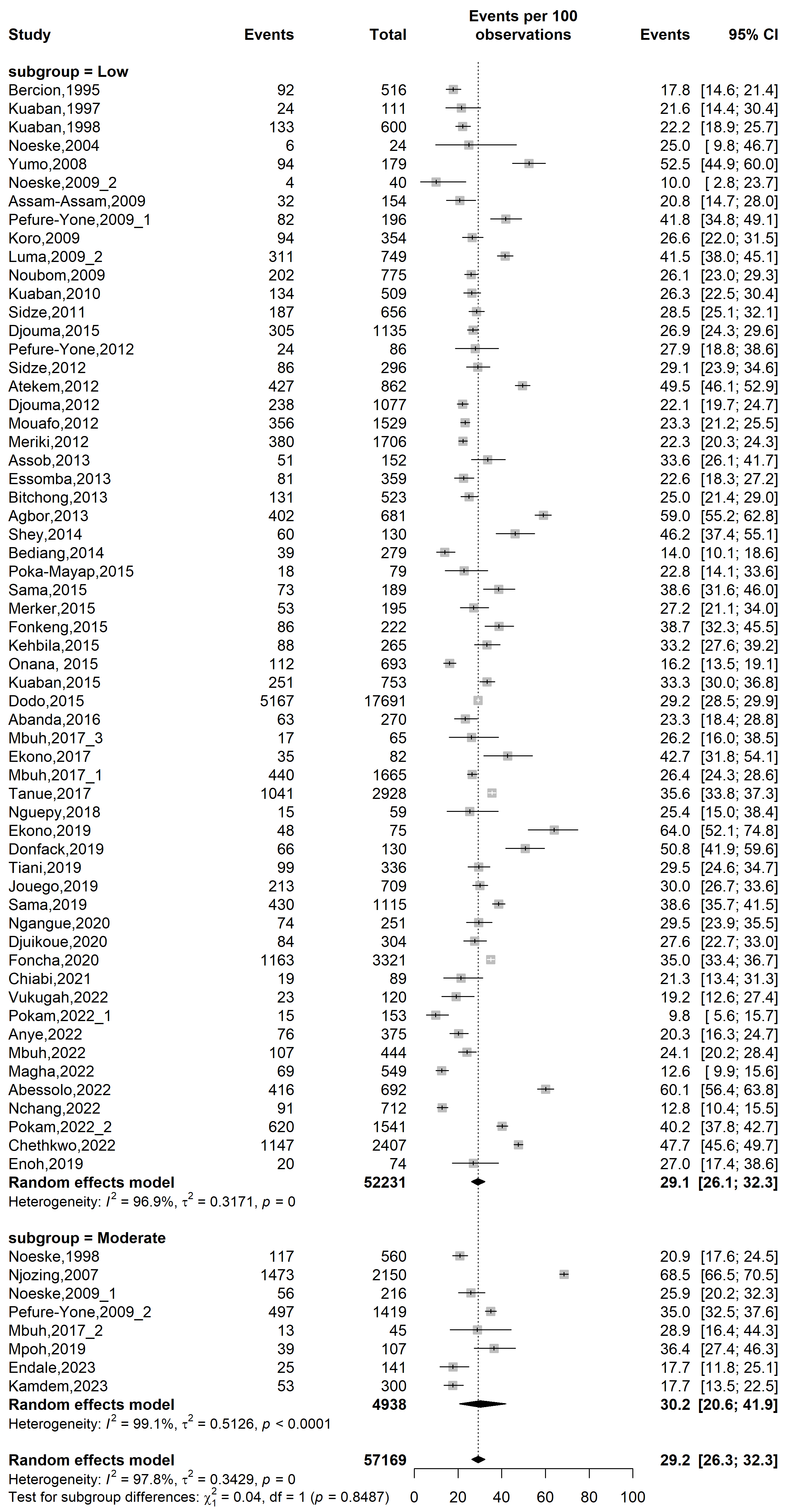

**Fig. S9** Forest plot displaying the subgroup meta-analysis of pooled prevalence of HIV prevalence among tuberculosis patients in Cameroon by risk of bias, 1995-2023

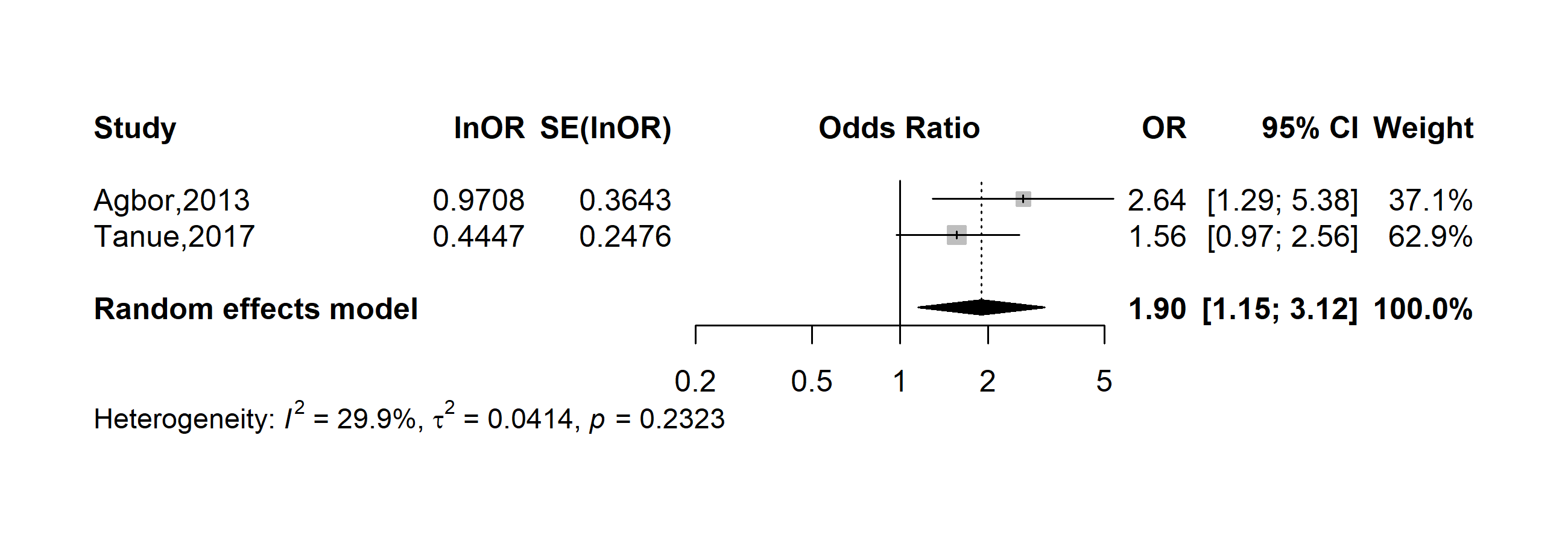

**Fig. S10** Pooled adjusted odds ratio of unfavorable outcome among human immunodeficiency virus and tuberculosis coinfected patients in Cameroon, 2013-2017 (Antiretroviral therapy: No vs. Yes)

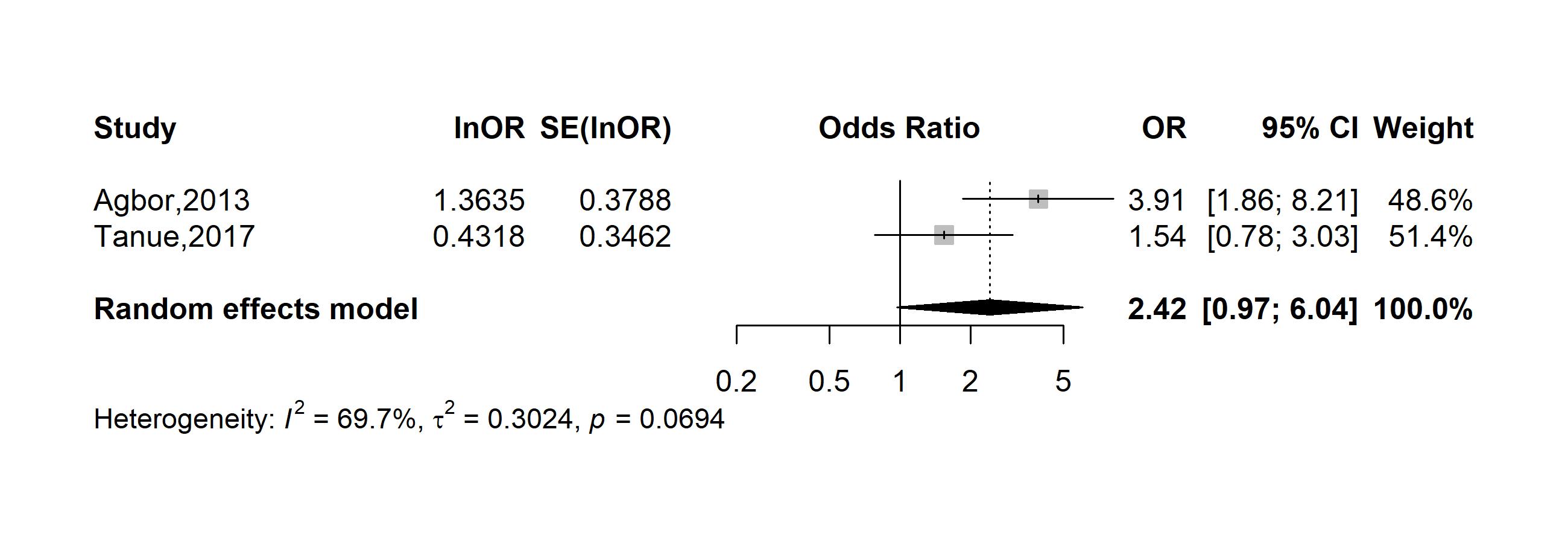

**Fig. S11** Pooled adjusted odds ratio of unfavorable outcome among human immunodeficiency virus and tuberculosis coinfected patients in Cameroon, 2013-2017 (Cotrimoxazole chemoprophylaxis: No vs. Yes)
